# The Effect of LLM Assistance on Diagnostic Accuracy: A Systematic Review and Meta-Analysis

**DOI:** 10.64898/2025.12.11.25341476

**Authors:** Isabel Tretow, Ege Ergon, Moritz Schwebel, Theresa Treffers, Isabell M. Welpe, Stefan Feuerriegel

## Abstract

Large language models (LLMs) are increasingly used in clinical settings, yet their effect on diagnostic accuracy of physicians has not been systematically quantified. We conducted a systematic review and meta-analysis of peer-reviewed articles and preprints published between January 2020 and June 2026 comparing diagnostic accuracy of physicians with and without LLM assistance. Eligible studies were randomized or non-randomized comparative studies using parallel-group or within-subject designs. We assessed potential risk of bias via PROBAST. Across 42 studies and 101 effect sizes, LLM assistance was associated with a small improvement in diagnostic accuracy (Hedges’ *g* = 0.22, 95% CI [0.18, 0.27], *p* < 0.001). However, the magnitude of the improvement varied substantially across subgroups (e.g., across LLMs or medical fields). These findings show that LLMs can improve the diagnostic accuracy of physicians, but conditions for successful LLM assistance remain unclear. Further clinical evidence is needed to guide safe and effective integration into practice.

## 1 Introduction

Accurate medical diagnosis is crucial for patient safety and effective treatment planning^1^. Still, diagnostic errors are common, with large risks to patient health. In the US alone, nearly 800,000 patients die or are left with a permanent disability each year due to diagnostic errors^2^. Reducing diagnostic errors remains an ongoing challenge in achieving diagnostic excellence. Large language models (LLMs), a form of artificial intelligence that can generate human-like outputs^3^, may help address this challenge by improving diagnostic accuracy and coming closer to diagnostic excellence. Unlike traditional rule-based or statistical clinical decision support systems (CDSS) (see Supplementary Table S1 for a detailed definition), LLMs generate natural-language responses based on pretrained language models and have shown strong performance across medical decisions: For instance, LLMs can assess health conditions, support in developing treatment strategies, or are able to solve challenging diagnostic cases^4–7^.

However, the benefit of LLM assistance in medical decision-making, specifically, diagnostic accuracy, remains unclear. Many studies compare the diagnostic accuracy of stand-alone LLMs with that of physicians^8^; however, in medical practice, LLMs are unlikely to operate independently and will more likely function under physician supervision by serving as assistive tools rather than replacements. Still, the benefits of such assistance appear mixed, and LLM assistance may help or even hurt. On the one hand, LLMs can leverage their vast amount of training data, including cases on rare diseases, to provide diagnostic evidence beyond the experience of an individual physician^9^. For instance, ChatGPT was able to provide the correct diagnosis for a young boy after 17 physicians had failed to do so^4^. Additionally, LLMs may complement human reasoning by providing alternative diagnostic perspectives. Thereby, LLMs may help overcome human cognitive biases such as confirmation and anchoring bias^10,11^. On the other hand, LLMs are at risk of producing misinformation due to limited common-sense reasoning or hallucinations (i.e., plausible, but factually incorrect responses, resulting from training limitations and the probabilistic nature of machine learning)^3,12,13^. For example, a patient developed bromism after following advice from ChatGPT to substitute sodium chloride with sodium bromide^14^. Such cases highlight the potential harm of LLMs, thereby emphasizing that responses from LLMs have to be critically reviewed and contextualized^8,15^. Physicians may still inadvertently follow wrong LLM advice, which poses new risks for diagnostic errors. While individual studies have examined the effect of LLM assistance on the diagnostic accuracy of physicians, systematic evidence remains limited, with two recent meta-analyses summarizing the effect of LLM assistance across only two^16^ and nine studies^17^. Given the fast-paced research on medical LLMs, a broader, up-to-date evidence synthesis is needed to quantify the magnitude and variability of LLM assistance on physicians’ diagnostic accuracy across clinical and study contexts.

To fill this gap, we conducted a meta-analysis on the effect of LLM assistance on the diagnostic accuracy of physicians. Our results show a significant, positive effect of LLM assistance on diagnostic accuracy. We further identify key determinants of effective collaboration, such as the LLM model, the medical field, the career stage, and the response format, and also identify areas in need of more research.

## 2 Results

### 2.1 Study selection and characteristics

#### Overall study design and outcome characteristics

Our systematic review and meta-analysis included 42 studies (Table 1), contributing *k* = 101 effect sizes. Across these studies, more than 680 physicians participated and evaluated more than 11,000 cases (percentages below refer to the proportion of studies rather than effect sizes). Top-1 accuracy was the most common accuracy measure (38/42, 90.5%), followed by top-3 accuracy (26.2%); top-2 and top-10 accuracy were each assessed in one study (2.4%). Overall, nine studies (21.4%) reported more than one accuracy measure. The majority of studies assessed diagnostic accuracy using binary outcomes (90.5%), whereas four studies (9.5%) used continuous outcomes.

**Table 1.** Characteristics of studies included in the meta-analysis (*n* = 42). *Note.* Accuracy measure: measure used to determine diagnostic accuracy (top-1, top-2, top-3, or top-10 accuracy); Outcome type: form of the extracted diagnostic accuracy outcome (binary, continuous); LLM model: specific model used in the study (e.g., GPT-4, Articulate Medical Intelligence Explorer (AMIE), GPT-4o); Task format: type of diagnostic task (open-ended, multiple choice, classification); Input form: format of patient information provided to the LLM (image, text, or multimodal [combination of text- and image-based data]); Response format: style of the model’s reasoning or output (Chain-of-Thought (CoT): step-wise explanations; differential: list of differential diagnoses; free text: open-ended answers to prompts, e.g., image descriptions); Customized prompt: indicates whether physicians could customize prompts or ask follow-up questions (yes, no); Medical field: main clinical domain of the cases, fields coded as “Other” are specified in parentheses; Career stage: seniority or training status of participating physicians (residents, attendings, mixed); Setting of patient cases: type of diagnostic cases used in the study (published cases: publicly available cases that could have been part of LLM training data; real-world cases: non-public cases, e.g., from a hospital; adapted real-world cases: non-public cases with additional modifications); Design: type of study design (within-subject, parallel-group); Publication status: peer-review status of the study at the time of dissemination; Country: geographic or national context in which study participants were recruited; No. of physicians: number of physicians participating in the study; No. of cases: number of unique cases within the respective study; No. of case observations: total number of physician-case examinations.

| Citation | Author | Accuracy measure | Outcome type | LLM model | Task format | Input form | Response format | Customized prompt | Medical field | Career stage | Setting of patient cases | Design | Publication status | Country | No. of physicians | No. of cases | No. of case observations |
| --- | --- | --- | --- | --- | --- | --- | --- | --- | --- | --- | --- | --- | --- | --- | --- | --- | --- |
| 19 | Atakır et al. | Top-1 | Binary | GPT-4o | Multiple choice | Multimodal, Text | Free text | No | Radiology | Resident | Published cases | Within-Subject | Peer-reviewed | Türkiye | 9 | 129 | 2,322 |
| 20 | Çamur et al. | Top-1 | Binary | GPT-5 | Open-ended | Multimodal | Differential | No | Radiology | Attending | Published cases | Within-Subject | Peer-reviewed | Türkiye | 2 | 86 | 516 |
| 21 | Cesur et al. | Top-1 | Binary | GPT-4o | Open-ended | Text | Differential | No | Radiology | Attending | Published cases | Within-Subject | Peer-reviewed | Türkiye | 4 | 80 | 640 |
| 22 | Chen et al. | Top-1 | Binary | ClinDiag-GPT | Open-ended | Text | Differential, free text | Yes | General medicine | Resident | Published cases | Parallel-group | Peer-reviewed | China | 6 | 60 | 120 |
| 23 | Dezoteux et al. | Top-1 | Binary | GPT-o1 | Multiple choice | Multimodal | CoT | No | Other (Dermatology) | Mixed | Published cases | Within-Subject | Peer-reviewed | France | 13 | 50 | 1,300 |
| 24 | Goh et al. | Top-1 | Binary | GPT-4 | Open-ended | Text | Free text | Yes | General medicine | Mixed | Real-world cases | Parallel-group | Peer-reviewed | United States | 50 | 6 | 244 |
| 25 | Healy et al. | Top-1 | Continuous | GPT-4o | Open-ended | Text | Free text | Yes | General medicine | Mixed | Real-world cases | Within-Subject | Peer-reviewed | United Kingdom | 22 | 4 | 87 |
| 26 | Horiuchi et al. | Top-1, Top-3 | Binary | GPT-4 | Open-ended | Text | CoT | No | Radiology | Resident, attending | Published cases | Within-Subject | Peer-reviewed | Japan | 2 | 106 | 424 |
| 27 | Ji et al. | Top-1 | Binary | SpAgents | Classification | Multimodal | CoT | Yes | Other (Rheumatology) | Resident, attending | Real-world cases | Within-Subject | Peer-reviewed | China | 7 | 100 | 2,100 |
| 28 | Jin et al. | Top-1 | Binary | GPT-4V | Classification | Image | CoT | No | Radiology | Resident, attending | Real-world cases | Within-Subject | Peer-reviewed | China | 6 | 103 | 1,236 |
| 15 | Kim et al. | Top-1, Top-3 | Binary | GPT-4 | Open-ended | Text | Differential | Yes | Radiology | Resident | Real-world cases | Within-Subject | Peer-reviewed | Germany | 6 | 40 | 228 |
| 29 | Kim et al. | Top-3 | Binary | GPT-4-Turbo | Open-ended | Multimodal | Differential | Yes | Radiology | Resident | Published cases | Within-Subject | Peer-reviewed | Germany | 9 | 30 | 262 |
| 30 | Kowshik et al. | Top-1 | Binary | LUNAR | Multiple choice | Text | CoT | No | Other (Neurology) | Attending | Real-world cases | Within-Subject | Preprint | Not reported | 12 | 100 | 1,156 |
| 31 | Le Guellec et al. | Top-3 | Binary | Gemini 1.5 Pro | Open-ended | Multimodal | Differential, free text | No | Radiology | Attending | Published cases | Within-Subject | Peer-reviewed | France | 3 | 53 | 318 |
| 32 | Liu et al. | Top-1 | Binary | MedFound-DX-PA | Open-ended | Text | CoT | No | General medicine | Resident, attending | Real-world cases | Within-Subject | Peer-reviewed | China | 12 | 300 | 3,600 |
| 33 | Long et al. | Top-3 | Binary | GPT-4o | Open-ended | Text | Differential | No | General medicine | Resident, attending | Real-world cases | Within-Subject | Peer-reviewed | China | 6 | 60 | 1,440 |
| 34 | McDuff et al. | Top-1, Top-10 | Binary | AMIE | Open-ended | Text | Differential, free text | Yes | General medicine, other (neurology, paediatrics, psychiatry) | Attending | Published cases | Parallel-group | Peer-reviewed | United States | 20 | 244 | 488 |
| 35 | Mukherjee et al. | Top-1 | Binary | GPT-4V | Multiple choice | Multimodal | Free text | No | Radiology | Resident, attending | Published cases | Within-Subject | Peer-reviewed | United States | 6 | 72 | 864 |
| 36 | Oon et al. | Top-1 | Continuous | GPT-4 | Open-ended | Text | Free text | Yes | Other (Pathology) | Mixed | Published cases | Within-Subject | Peer-reviewed | Singapore | 9 | 10 | 180 |
| 37 | Qazi et al. | Top-1 | Continuous | GPT-4o | Open-ended | Text | Free text | Yes | General medicine | Attending | Adapted real-world cases | Parallel-group | Peer-reviewed | Pakistan | 58 | 6 | 342 |
| 38 | Schramm et al. | Top-1, Top-3 | Binary | GPT-4.1 | Open-ended | Text | Differential | No | Radiology | Resident, attending | Published cases | Within-Subject | Peer-reviewed | Germany | 12 | 40 | 942 |
| 39 | Sheng et al. | Top-1, Top-3 | Binary | GPT-4o | Open-ended | Text | Differential | No | Radiology | Resident, attending | Real-world cases | Within-Subject | Peer-reviewed | China | 6 | 228 | 2,736 |
| 40 | Siepmann et al. | Top-1, Top-3 | Binary | GPT-4 | Open-ended | Text | Differential, free text | Yes | Radiology | Resident | Real-world cases | Within-Subject | Peer-reviewed | Germany | 6 | 40 | 480 |
| 41 | Song et al. | Top-1 | Binary | Gemini 3.0 Pro | Multiple choice | Text | Differential, free text | No | Radiology | Resident, attending | Published cases | Within-Subject | Peer-reviewed | South Korea | 10 | 93 | 1,860 |
| 42 | Song et al. | Top-1, Top-3 | Binary | Claude 3.5 Sonnet | Open-ended | Multimodal | Differential | No | Radiology | Resident, attending | Real-world cases | Within-Subject | Peer-reviewed | South Korea | 5 | 127 | 1,270 |
| 18 | Spitzer et al. | Top-1 | Binary | GPT-4 | Open-ended | Multimodal | CoT, Differential, free text | No | Radiology | Attending | Published cases | Parallel-group | Peer-reviewed | United States | 101 | 20 | 2,020 |
| 43 | Ver Berne et al. | Top-1 | Binary | GPT-4 | Open-ended | Text | Differential | No | Radiology | Mixed | Real-world cases | Within-Subject | Peer-reviewed | Belgium | 6 | 25 | 300 |
| 44 | Wang et al. | Top-1 | Binary | GPT-4o | Classification | Multimodal | Free text | No | Radiology | Resident | Real-world cases | Within-Subject | Peer-reviewed | China | 5 | 77 | 770 |
| 45 | Wang et al. | Top-1 | Continuous | PsychFound | Open-ended | Text | CoT | Yes | Other (Psychiatry) | Resident | Real-world cases | Parallel-group | Peer-reviewed | China | 24 | 200 | 200 |
| 46 | Wang et al. | Top-1 | Binary | DeepSeek-R1 | Open-ended | Text | CoT | No | Radiology | Resident, attending | Real-world cases | Within-Subject | Peer-reviewed | China | 6 | 500 | 6,000 |
| 47 | Wang et al. | Top-1 | Binary | RetiZero | Multiple choice | Image | Differential | No | Ophthalmology | Resident, attending | Real-world cases | Within-Subject | Peer-reviewed | Singapore, United States, China | 19 | 104 | 3,952 |
| 48 | Wang et al. | Top-1 | Binary | GPT-4 | Classification | Text | Free text | No | Radiology | Resident, attending | Real-world cases | Parallel-group | Peer-reviewed | China | 30 | 159 | 357 |
| 49 | Weuthen et al. | Top-1 | Binary | GPT-4 | Multiple choice | Text | Free text | Yes | General medicine | Mixed | Adapted real-world cases | Parallel-group | Peer-reviewed | Germany | 32 | 1 | 32 |
| 50 | Wu et al. | Top-1 | Binary | DeepSeek-R1 | Open-ended | Text | Differential | No | General medicine | Resident | Published cases | Parallel-group | Peer-reviewed | China | 32 | 48 | 96 |
| 51 | Wu et al. | Top-1, Top-2 | Binary | EyeFM | Open-ended, Classification | Multimodal, Image | Differential, free text | No | Ophthalmology | Resident, attending | Real-world cases | Within-Subject, Parallel-group | Peer-reviewed | China | 26 | 2,106 | 2,546 |
| 52 | Xu et al. | Top-1 | Binary | GPT-4o | Classification | Image | Free text | No | Radiology | Resident, attending | Real-world cases | Within-Subject | Peer-reviewed | China | 6 | 110 | 1,320 |
| 53 | Yan et al. | Top-1, Top-3 | Binary | PanDerm | Open-ended, Multiple choice | Image | Differential | No | Other (Dermatology) | Mixed | Published cases | Within-Subject | Peer-reviewed | Multiple countries | 78 | 1,711 | 4,662 |
| 54 | Yao et al. | Top-1 | Binary | ThyGPT | Classification | Image | Free text | Yes | Radiology | Resident, attending | Real-world cases | Within-Subject | Peer-reviewed | China | 6 | 3,376 | 40,512 |
| 55 | Zhang et al. | Top-1 | Binary | GPT-4o | Classification | Multimodal | Free text | No | Radiology | Resident, attending | Real-world cases | Within-Subject | Peer-reviewed | China | 4 | 454 | 1,816 |
| 56 | Zhou et al. | Top-1 | Binary | ConfiDx (LLaMA-3.1-70B) | Open-ended | Text | Free text | No | Other (Cardiology, endocrinology) | Attending | Adapted real-world cases | Within-Subject | Peer-reviewed | United States | $\geq 4$ | 200 | 800 |
| 57 | Zhou et al. | Top-3 | Binary | HeartAgent (Llama-3.3-70B) | Open-ended | Multimodal | Differential, free text | Yes | Other (Cardiology) | Attending | Adapted real-world cases | Within-Subject | Preprint | United States | 2 | 100 | 200 |
| 58 | Zhou et al. | Top-1 | Binary | EpiScreen | Classification | Text | Free text | No | Other (Neurology) | Resident, attending | Adapted real-world cases | Within-Subject | Preprint | United States | 4 | 300 | 1,200 |

### LLM and interaction characteristics

The studies evaluated 24 distinct LLMs. GPT-4 and GPT-4o were the most frequently studied models (21.4% each), followed by GPT-4V and DeepSeek-R1 (4.8% each); all remaining models were evaluated in one study each. Diagnostic tasks were most commonly open-ended (64.3%), followed by classification tasks (21.4%; e.g., differentiating between malignant and benign tumors) and multiple-choice tasks (19.0%), including two studies that evaluated more than one task format. Regarding the input form (i.e., how case information was provided to the LLM), text-only input was evaluated in 25 studies (59.5%), multimodal (text-and-image) input in 13 (31.0%), and image-only input in six (14.3%); two studies evaluated more than one input form. LLM response formats varied, with free-text responses evaluated in 22 studies (52.4%), differential diagnoses in 20 (47.6%), and Chain-of-Thought (CoT; i.e., referring to an LLM response that includes a detailed, step-by-step reasoning^18^) responses in nine (21.4%); eight studies (19.0%) evaluated more than one response format. Physicians were allowed to customize prompts or ask follow-up questions in 14 studies (33.3%), whereas prompts were not physician-customizable in 28 studies (66.7%).

### Clinical and study characteristics

Radiology was the most frequently studied medical field (52.4%), while general medicine was represented in nine studies (21.4%). The remaining studies covered a broad range of specialties, including ophthalmology, dermatology, neurology, rheumatology, pathology, psychiatry, and cardiology. Residents were represented in 26 studies (61.9%) and attendings in 27 (64.3%), including 18 studies (42.9%) that included both career stages; seven studies (16.7%) reported mixed-career samples. Real-world cases were evaluated in 21 studies (50.0%), published cases in 16 (38.1%), and adapted real-world cases in five (11.9%). 32 studies (76.2%) used within-subject comparisons, nine (21.4%) used parallel-group designs, and one study (2.4%) included both designs. Finally, 39 studies (92.9%) were peer-reviewed, and three (7.1%) were preprints.

### 2.2 Meta-analysis

#### Overall effect and accuracy measures

Across 101 effect sizes, LLM assistance showed a small positive effect on physicians’ diagnostic accuracy (Hedges’ *g* = 0.22, 95% CI [0.18, 0.27], *p* < 0.001; Figure 1). Heterogeneity was substantial (*I*^2^ = 73.12%; total *τ* ^2^ = 0.02). Estimates were similar for top-1 (*g* = 0.22, 95% CI [0.17, 0.26]) and top-3 accuracy (*g* = 0.25, 95% CI [0.17, 0.33]). Estimates for top-2 (*g* = 0.22, 95% CI [−0.02, 0.45]) and top-10 accuracy (*g* = 0.21, 95% CI [−0.40, 0.83]) were based on only two effect sizes each and were therefore estimated with considerable uncertainty (i.e., had wider confidence intervals; Figure 2).

**Fig. 1.**
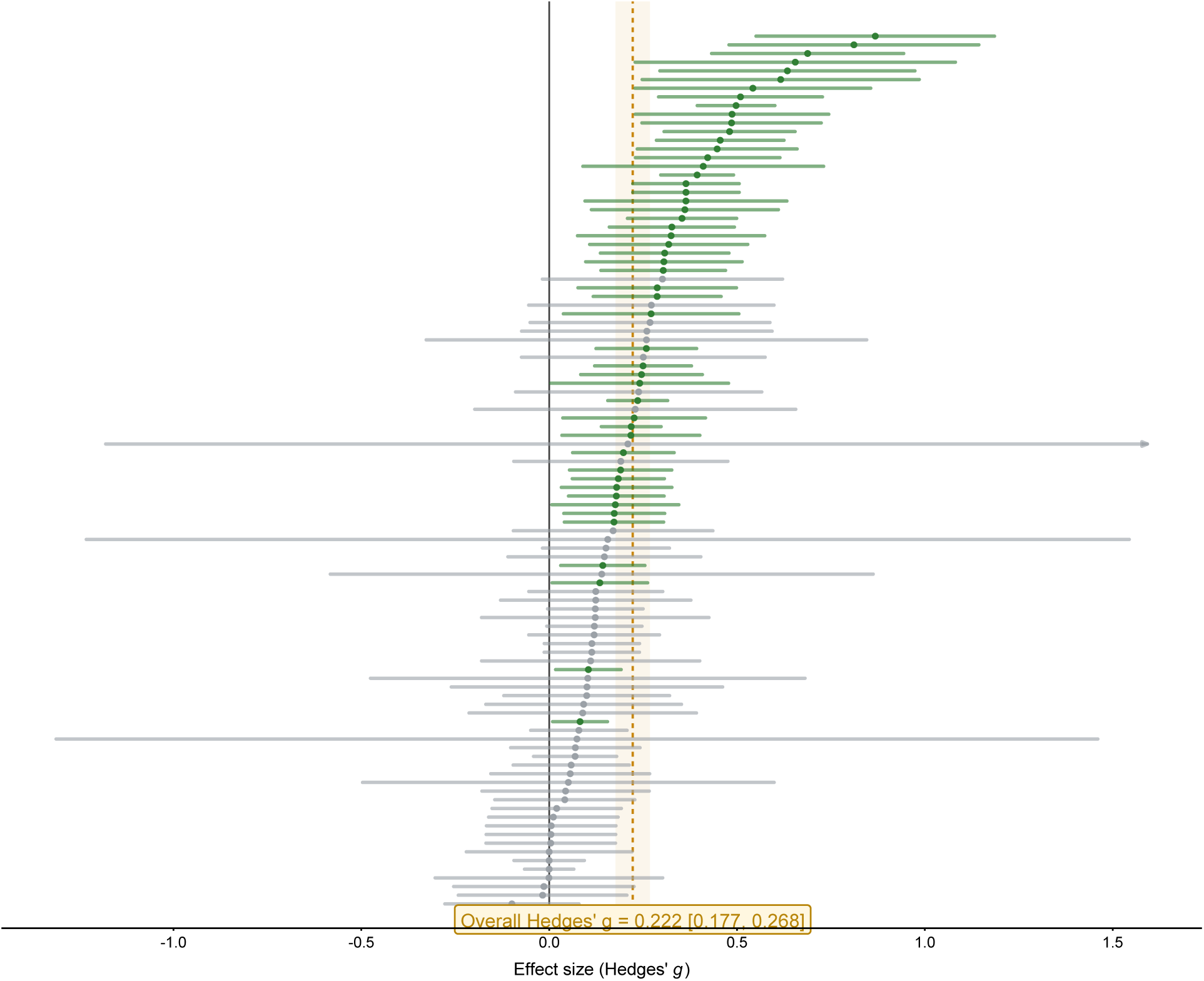
Forest plot of effect sizes (*k* = 101) comparing diagnostic accuracy with and without LLM assistance. Each horizontal line represents the 95% CI for an individual effect size, with the central marker indicating Hedges’ *g*. The pooled effect size (Hedges’ *g* = 0.22, 95% CI [0.18, 0.27]) is indicated by the dashed vertical line and yellow-shaded confidence band. The gray line corresponds to an effect size of *g* = 0. A detailed forest plot including individual effect size labels and weights is provided in Supplementary Figure S3.

**Fig. 2.**
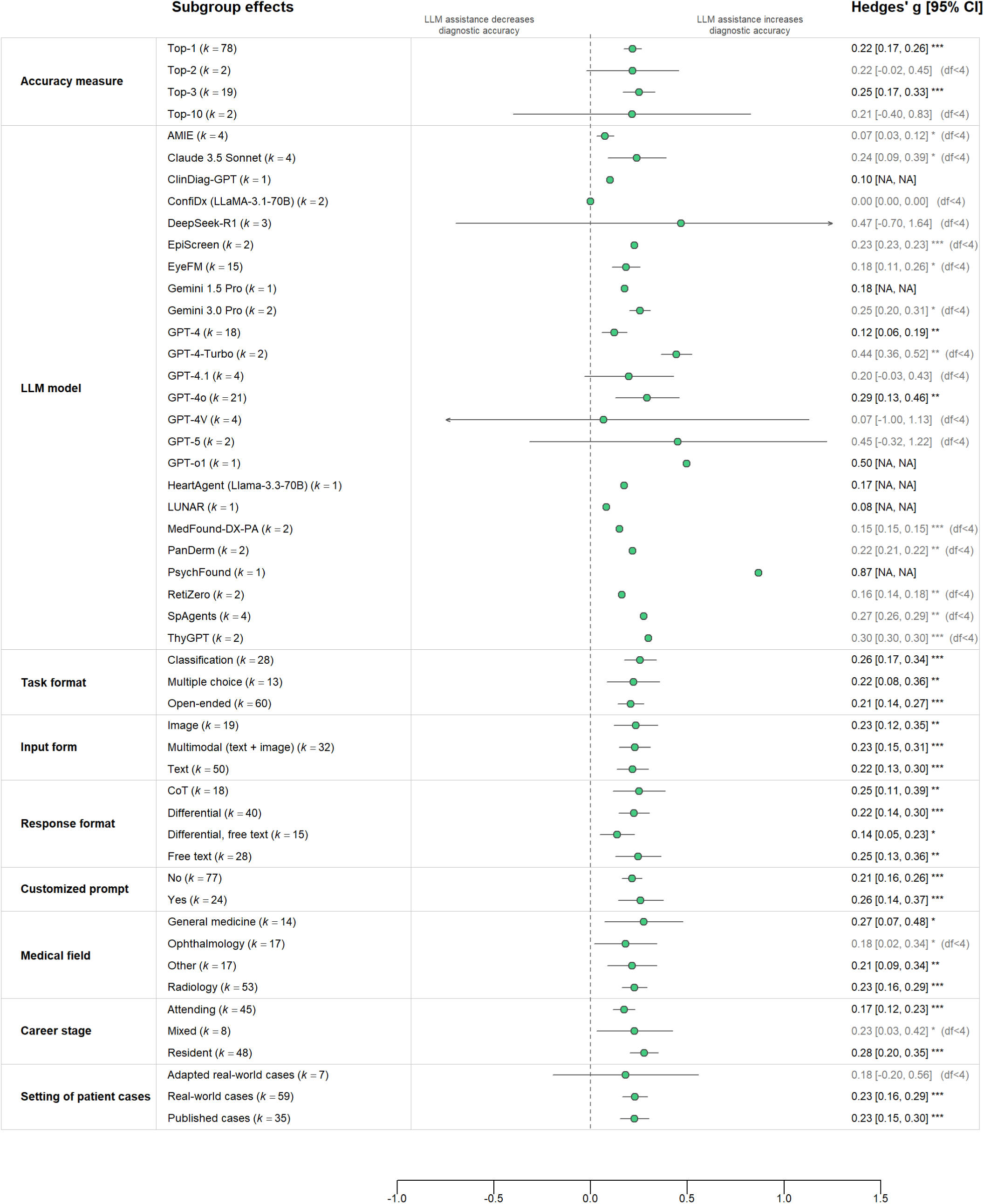
Subgroup effect sizes by moderator. Each point shows the pooled Hedges’ *g* for one sub-group, estimated in a separate meta-regression for the respective moderator; horizontal lines indicate 95% CIs. Models were fitted without an intercept so that each estimate directly represents the pooled effect for the corresponding subgroup. Standard errors, 95% CIs, and *p*-values were calculated using cluster-robust variance estimation (CR2) with Satterthwaite degrees of freedom, clustered at the paper/case-corpus level. *p*-values test whether the effect within each subgroup differs from zero; differences between subgroups were assessed separately using omnibus tests (Supplementary Table S2). Here, *k* (shown in brackets behind each subgroup) denotes the number of effect sizes included in each subgroup (when *k* = 1, no CI is available (labeled as NA)). For descriptions of the moderators, see Table 1. The vertical dashed line marks *g* = 0, indicating no difference in diagnostic accuracy between physicians with and without LLM assistance. Significance levels: * *p* < 0.05, ** *p* < 0.01, *** *p* < 0.001. Labels “*df* < 4” indicate subgroup estimates based on few independent study clusters. CoT refers to Chain-of-Thought.

### LLM and interaction characteristics

For the most frequently evaluated GPT variants, estimates were *g* = 0.12 (95% CI [0.06, 0.19]) for GPT-4 and *g* = 0.29 (95% CI [0.13, 0.46]) for GPT-4o. Several other (domain-specific) estimates were based on only a small number of effect sizes, which limits comparisons across models. Across task formats, estimates were similar for open-ended and multiple-choice tasks (*g* = 0.21, 95% CI [0.14, 0.27] and *g* = 0.22, 95% CI [0.08, 0.36], respectively), and slightly higher for classification tasks (*g* = 0.26, 95% CI [0.17, 0.34]). Positive estimates were also observed across input forms, with similar estimates for text (*g* = 0.22, 95% CI [0.13, 0.30]), image (*g* = 0.23, 95% CI [0.12, 0.35]), and multimodal input (*g* = 0.23, 95% CI [0.15, 0.31]). Across LLM response formats, estimates ranged from *g* = 0.14 (95% CI [0.05, 0.23]) for combined differential and free-text responses to *g* = 0.25 for free-text (95% CI [0.13, 0.36]) and CoT responses (95% CI [0.11, 0.39]). Estimates were similar when physicians could customize prompts (*g* = 0.26, 95% CI [0.14, 0.37]) or not (*g* = 0.21, 95% CI [0.16, 0.26]).

### Clinical and study characteristics

Subgroup estimates varied across medical fields; for example, *g* = 0.18 (95% CI [0.02, 0.34]) was observed for ophthalmology in comparison to *g* = 0.27 (95% CI [0.07, 0.48]) for general medicine. Estimates were higher for residents (*g* = 0.28, 95% CI [0.20, 0.35]) than for attendings (*g* = 0.17, 95% CI [0.12, 0.23]), with mixed-career groups yielding an estimate of *g* = 0.23 (95% CI [0.03, 0.42]). Estimates were similar for real-world (*g* = 0.23, 95% CI [0.16, 0.29]) and published cases (*g* = 0.23, 95% CI [0.15, 0.30]); the estimate for adapted real-world cases was less precise (*g* = 0.18, 95% CI [−0.20, 0.56]). The moderator analyses and a meta-regression combining all moderators in a single model are reported in Supplementary Tables S2 and S3.

### 2.3 Risk-of-bias assessment

To assess the risk of bias, we used the Prediction Model Risk of Bias Assessment Tool (PROBAST). Overall, we conducted 45 risk-of-bias assessments and classified 32 (71%) as high risk of bias and six (13%) as having high applicability concerns (see Supplementary Figure S1 for details). Assessments were generally conducted at the study level. However, for two articles reporting multiple distinct experiments^51,53^, experiments were assessed separately because the underlying risk-of-bias profiles were different. We assigned a high risk of bias when the same physicians evaluated the same cases with and without LLM assistance (within-subject design) or when test data could have been included in LLM training data (e.g., in the case of publicly available cases). Applicability concerns were mostly assigned for very specific physician samples (e.g., radiologists with *<* 6 months of experience). A sensitivity analysis restricted to studies with low risk of bias yielded a similarly positive effect of LLM assistance on diagnostic accuracy (Hedges’ *g* = 0.28, 95% CI [0.12, 0.43]; Supplementary Figure S2).

### 2.4 Robustness and sensitivity checks

Sensitivity analyses supported the stability of the pooled effect. We assessed potential small-study effects using a funnel plot (see Supplementary Figure S4) and a CR2-robust Egger-type regression^59^ with a sample-size-based precision proxy. We found no evidence of small-study effects (*β* = −0.28, *p* = 0.52).

Results were similar when a cluster-level random intercept was added, with studies using over-lapping patient cases grouped into a common cluster. This yielded a four-level meta-analytic model comprising sampling error, effect sizes, experiments, and paper/case-corpus clusters, while cluster-robust inference remained clustered at the paper/case-corpus level (*g* = 0.23, 95% CI [0.18, 0.29], *p* < 0.001; 40 clusters, 101 effect sizes). The pooled estimate also remained robust when varying assumptions about within-subject correlation (*r* = 0 – 0.9: *g* = 0.222 – 0.223) and correlation between case evaluations from the same physician (intraclass correlation coefficient, ICC = 0 – 0.20: *g* = 0.21 – 0.23; details in Supplementary Table S4). Leave-one-out analyses further supported the stability of the pooled effect: excluding individual effect sizes, experiments, or studies yielded estimates between *g* = 0.22 and *g* = 0.23, with all *p* < 0.001 (Supplementary Tables S6, S7, and S8).

## 3 Discussion

In our systematic review and meta-analysis, we analyzed the effect of LLM assistance on the diagnostic accuracy of physicians. We initially identified 6,721 records, from which 42 studies were ultimately included in our meta-analysis. We found a statistically significant but small positive effect of LLM assistance on the diagnostic accuracy of physicians (Hedges’ *g* = 0.22, 95% CI [0.18, 0.27], *p* < 0.001). However, the magnitude of the effect varied across studies and settings, indicating that the benefits of LLM assistance are not uniform. These findings are particularly relevant given the increasing use of LLMs in clinical practice: among physicians who use LLMs, around 76% report using them for clinical decisions^60^. While LLMs have shown strong performance across medical use cases^6,61–63^, LLM use also involves inherent risks, particularly as hallucinations and inappropriate reliance on model outputs may threaten patient safety^12^.

Our results extend prior work that has assessed the stand-alone diagnostic accuracy of LLMs in comparison to physicians^5,8,64^. In contrast, we focus on scenarios where physicians use LLMs as collaborative aids rather than in full automation. In clinical practice, it is unlikely that LLMs will make diagnostic decisions in an automated manner without human oversight; rather, physicians will be supported by LLMs and may consult LLMs as assistance to obtain diagnostic suggestions, compare reasoning, or refine their decisions^60^. While such collaborations can improve diagnostic accuracy, they are also prone to cognitive risks. Physicians may over-rely on model suggestions (automation bias) or anchor on an initial LLM-generated diagnosis, shaping subsequent reasoning^65–67^. LLMs producing inaccurate or hallucinatory information can also compromise patient safety. Therefore, effective integration of LLMs into clinical workflows requires understanding how physicians use LLM assistance and when collaboration is most beneficial.

We find a positive effect of LLM assistance on the diagnostic accuracy of physicians, but the magnitude of this effect varies across studies, thereby underscoring that the effectiveness of LLM assistance depends on several determinants. We identify four key determinants of effective collaboration between physicians and LLMs: the underlying LLM model, the medical field, the physician’s career stage, and the response format used to present or solicit diagnostic input. These factors shape how physicians interpret, trust, and apply LLM recommendations in clinical reasoning, but more research is needed to effectively integrate LLMs in practice.

### LLM models

We found no systematic performance differences across LLMs, likely due to limited LLM diversity. Most studies assessed closed-source models such as GPT-4, while open-weight^32,46,50^ and reasoning models were rarely used^20,23,27,41,46,50^. Given the flexibility of open-weight models for customization^30,51,56,68,69^ and the potential of reasoning models to iteratively criticize and refine LLM outputs^70^, future work should assess the potential of fine-tuned open-weight LLMs for currently rarely analyzed medical fields and how reasoning LLMs can improve diagnostic accuracy compared to standard models.

### Medical field

LLM assistance improved diagnostic accuracy across different medical fields. Still, research concentrates on fields with well-defined outcomes (e.g., radiology), which may limit generalizability to more complex and less standardized settings such as acute or critical care^71^. Here, additional research in different clinical contexts is needed.

### Career stage

LLM assistance improved diagnostic accuracy for residents, attendings, and mixed groups. Although prior work suggested stronger benefits for residents with limited clinical experience^9^, our results show that attendings also benefit, likely because their experience allows better judgment of when to accept or override LLM recommendations. Hence, LLM use may differ with experience, but which warrants further study.

### Response format

Our results further show a positive effect on diagnostic accuracy across various response formats. Still, hallucinated explanations might mislead physicians. While physicians in practice can ask clarifying questions, only around one-third of the studies allowed follow-up questions. Hence, future research might develop and test customized LLMs that signal uncertainty or pose follow-up questions to make it easier for physicians to detect hallucinations.

This study has limitations, which open avenues for future research. First, the included studies varied considerably in their designs, clinical settings, LLMs, and diagnostic tasks, and evidence remained sparse for some subgroups. Moderator findings should therefore be interpreted cautiously, particularly where estimates were based on few effect sizes. Second, some studies used publicly available patient cases that may have been included in LLM training data, potentially inflating the apparent effectiveness of LLM assistance. Moreover, several within-subject studies asked the same physicians to assess the same cases without and subsequently with LLM assistance; while we accounted for the resulting statistical dependency, this does not remove the potential for learning or carryover effects. Third, reporting of exact LLM versions and physician characteristics was frequently incomplete, limiting reproducibility and the assessment of model- or participant-specific effects. Fourth, although our search covered multiple bibliographic databases and used a broad search strategy, relevant studies may still have been missed given the rapidly evolving terminology, models, and evidence base in this field. We also included non-peer-reviewed preprints to broaden the coverage of this rapidly evolving literature, even though these findings had not yet undergone formal peer review. Fifth, while the evidence base included both general-purpose and domain-specialized LLM systems, general-purpose models, particularly GPT-4 variants, remained more frequently studied, and most specialized models were evaluated in only a single study. This uneven evidence base limits model-specific inference and precludes conclusions about whether specialized medical LLMs systematically differ from general-purpose models in their effectiveness as diagnostic assistance. Lastly, the meta-analysis shows the effect on diagnostic accuracy and does not imply clinical readiness, especially given that many questions around the translation to clinical decision support are still unresolved^72^. On the one hand, other relevant outcomes in clinical practice (e.g., efficiency) are not covered by our search and will require future research. On the other hand, the majority of studies use clinical vignettes and are not validated in routine care; LLM use in clinical decision-making may introduce new risks (e.g., hallucination, bias, overreliance^73^) as well as practical barriers of regulatory, ethical, or technological nature that will need to be considered carefully.

Based on these limitations, we derive five directions for future research. First, the studies in our sample assessed diagnostic accuracy in controlled settings rather than in real-world clinical environments, so that ecological validity in routine care is often unclear. Under time pressure and stress, physicians may be less critical of LLM recommendations and more prone to follow incorrect advice. Future research should analyze the effect of LLM assistance in real-world settings and how to effectively integrate LLMs into clinical practice. Second, some studies included training measures for physicians that educated them in how to effectively use LLMs in medical practice^15,29,36,37,40,45^. As physicians gain experience in using LLMs, future research could conduct longitudinal studies on how familiarity, habituation, and routine use affect accuracy and trust in LLM assistance over time. Third, biases in LLMs (e.g., on gender, race, ethnicity) can negatively affect patient health^74–76^. However, no study in our sample analyzed whether LLM assistance increases such biases. Future research should assess whether LLM assistance promotes biases in clinical decision-making across patient groups. Fourth, clinical decision-making extends beyond diagnostic accuracy. For example, future research could include triage or treatment recommendations to establish a comprehensive understanding of LLM assistance in medicine. Fifth, evidence remains concentrated on general-purpose LLMs, while most domain-specialized LLM systems have been evaluated in only a single study. Future research should directly compare general-purpose and domain-specialized LLMs under comparable diagnostic tasks, physician populations, and assistance settings, and replicate such evaluations across clinical contexts. This would help determine whether domain specialization provides additional benefits for diagnostic accuracy of physicians, beyond those observed with general-purpose models.

In conclusion, LLMs can assist physicians in diagnosis, but it remains unclear under which conditions this assistance is most effective. Identifying these conditions is essential to maximizing patient outcomes and ensuring patient safety. Given the heterogeneous evidence base and limitations of the available studies, rigorous clinical evidence is needed to address open questions around real-world applicability and integration into practice.

## 4 Methods

### 4.1 Search strategy

We conducted our meta-analysis in line with the Preferred Reporting Items for Systematic Reviews and Meta-analyses (PRISMA)^77^ and pre-registered the protocol via PROSPERO^78^. A systematic search was performed in five databases: MEDLINE (accessed via the PubMed interface), Web of Science, EMBASE, Scopus, and medRxiv. Preprints were included intentionally, given the rapidly evolving nature of LLM technologies and the lag in peer-reviewed publication (for a separate sensitivity analysis excluding preprints see Supplementary Table S5). The search was restricted to English-language studies published between January 1, 2020, coinciding with the emergence of models such as GPT-3^79^, and June 26, 2026. Our literature search followed the PICO framework^80^: The population (P) included clinicians and medical professionals, the intervention (I) involved generating diagnoses with LLM assistance, the comparator (C) was a diagnosis without LLM assistance. The primary outcome (O) was diagnostic accuracy.

We developed our search string based on five conceptual blocks to identify studies examining LLM assistance of physicians in diagnostic settings. The first block comprised terminology referring to LLMs in general (e.g., “large language model*”, “generative artificial intelligence”) as well as specific model names (e.g., “GPT-4*”, “ChatGPT”, “AMIE” (Articulate Medical Intelligence Explorer)). The second block captured terms describing the interaction between physicians and LLMs, including broad descriptors such as “support”, “assist*”, or “collaborat*”, alongside more specific phrases such as “LLM-assist*” or “AI-augment*”. The third block focused on the professional population of interest, using terms such as “physician*” or “clinician*”, as well as a set of medical specialties derived from terms linked to the MeSH term “physicians” in PubMed. The fourth block focused specifically on the diagnostic task, including terms such as “diagnos*” or “clinical decision making”. A fifth block aimed to exclude publication types unlikely to contain primary empirical evidence, including reviews, meta-analyses, editorials, letters, and comments. The search strategy was developed iteratively, starting from search terms used in prior work comparing the stand-alone LLM performance with that of humans^8,81^ and a set of known eligible studies. We subsequently refined the search string until all known eligible studies were retrieved and further revised it based on feedback from five medical and methodological experts, including board-certified physicians and a research librarian. The complete search string, including database-specific adaptations, is provided in Appendix B.

### 4.2 Study selection and data extraction

Our initial search across MEDLINE, Web of Science, EMBASE, Scopus, and medRxiv yielded a total of 6,721 records including 2,671 duplicates. Two independent reviewers (I.T. and E.E.) first screened titles and abstracts and, in a second step, conducted a full-text screening. Any disagreements were resolved by discussion with a third reviewer (S.F.). The screening process was managed using Rayyan, a screening platform for systematic reviews^82^. Studies were included if they compared the diagnostic accuracy of physicians with and without LLM assistance. Diagnosis refers to identifying the disease entity of a patient, and accuracy refers to the proportion of correct diagnoses, measured as a top-*k* diagnosis and assessed as a binary outcome or on an accuracy-specific Likert scale (coded as “continuous” in Table 1). Inclusion criteria were independent of the clinical specialty, target population, disease, LLM used, or type of diagnostic assistance provided by the LLM. Studies were excluded if they: (1) did not assess a diagnostic task or did not report a diagnostic accuracy measure (e.g., they only staged, graded, or risk-stratified an already-established disease entity); (2) did not involve interaction between the physician and the LLM or the physician could not make the final diagnosis; or (3) evaluated an AI support system without an LLM component. Reviews, meta-analyses, editorials, letters, comments, conference abstracts, prospective study protocols, and qualitative studies were also excluded. After screening, we included 42 studies in our meta-analysis (details see flow diagram in Figure 3).

**Fig. 3.**
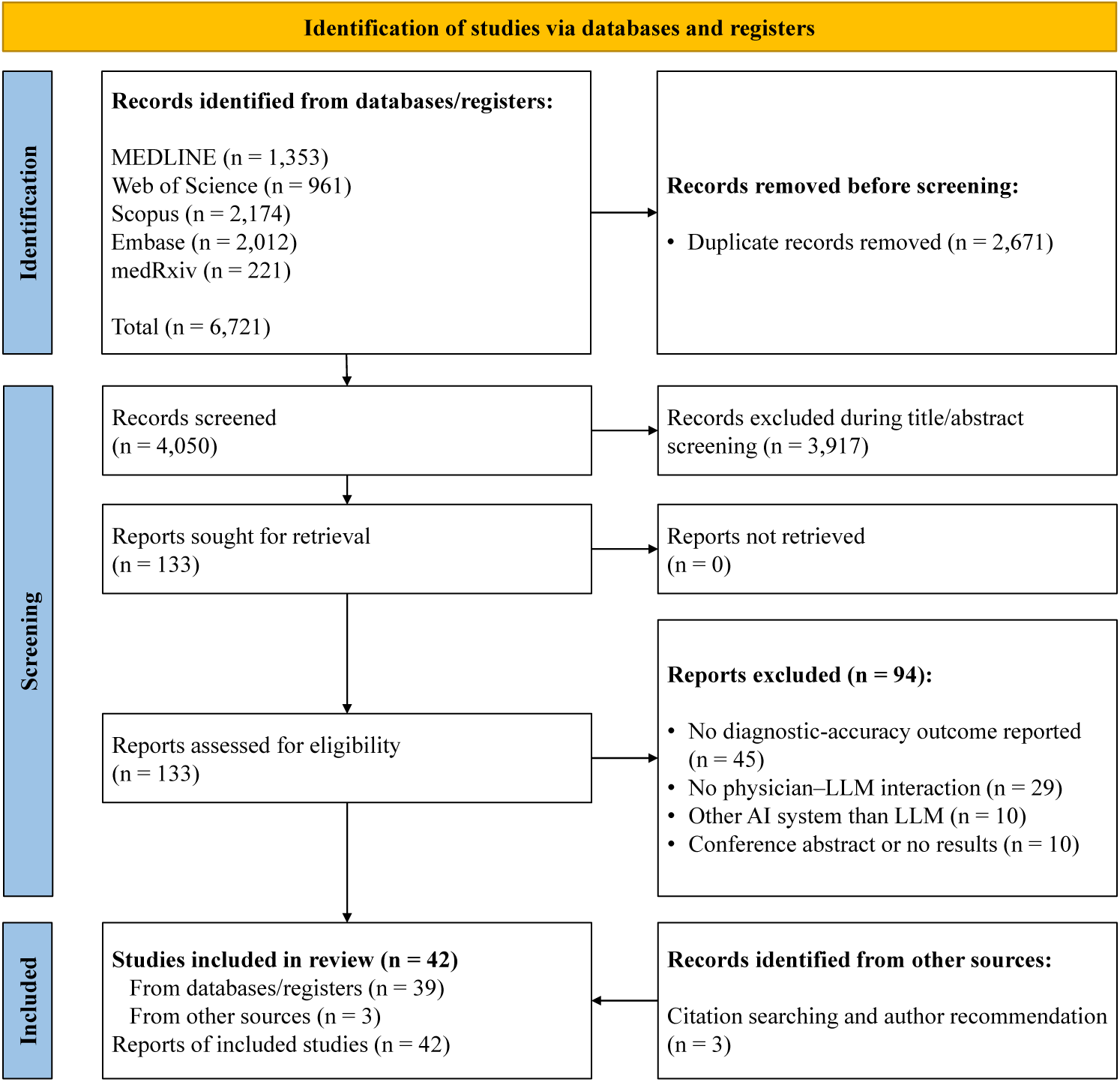
Flow diagram. The flow diagram summarizes our systematic review process, including the identification and screening of records.

Following best practice^8^, we used a predefined Excel sheet to extract data. Data extraction was conducted independently by two reviewers (I.T. and E.E.), who subsequently compared their coding and resolved discrepancies through discussion with a third reviewer (S.F.). Table 1 provides an overview of key dimensions extracted (additional explanations on dimensions in Appendix C). The complete coding scheme is available in our GitHub repository (see data availability). If studies did not report the statistics needed for effect size calculation, we reached out to the corresponding authors via e-mail and sent follow-up emails if no response was received. As a result, we received additional information for fifteen studies, which are included in our meta-analysis.

### 4.3 Risk-of-bias assessment

Risk of bias was assessed using PROBAST^83^, with two reviewers (I.T. and E.E.) independently conducting the assessment and resolving any disagreements through discussion with a third reviewer (S.F.). We excluded selected PROBAST questions that are specific to the evaluation of prediction studies and not applicable to our comparative analysis of physicians and LLM-assisted physicians. This involves all three questions from Domain 2 (predictors), as well as questions 3.3, 3.5, and 3.6 from Domain 3 (outcome) and questions 4.5 and 4.9 from Domain 4 (analysis)^8^. An overview of our PROBAST evaluation is available in Supplementary Figure S1.

### 4.4 Statistical analysis

The primary effect size metric for this analysis was Hedges’ *g* ^84^, referring to the difference in diagnostic accuracy of physicians with and without LLM assistance. Effect sizes and their sampling variances were calculated from the statistics reported in each study using design-appropriate standardized mean-difference estimators; detailed procedures for binary and continuous outcomes, within-subject designs, and within-physician clustering are provided in Appendix D.

While we coded at the granularity of reported studies (e.g., physician-level data), multiple granular estimates representing the same comparison and moderators were pooled before effect size calculation (a sensitivity analysis keeping the highest granularity for each study is also shown in Supplementary Figure S5). We applied a random-effects model given the expected heterogeneity across studies^80^. As several studies reported multiple diagnostic accuracy measures (see Table 1), effect sizes were statistically dependent, violating the independence assumption of many meta-analytic models. Thus, we employed a three-level meta-analytic model using the restricted maximum-likelihood (REML) estimation method^85^, nesting effect sizes within experiments, to account for dependencies in the data^81,86^. Specifically, we incorporated sampling variance at the effect size level (first level), variance between effect sizes of the same experiment (second level), and variance across experiments (third level). This approach allows inclusion of more than one accuracy measure per study^81,86^.

Standard errors, confidence intervals (CI), and *p*-values were obtained using cluster-robust variance estimation (CR2) with Satterthwaite degrees of freedom, clustering at the paper/case-corpus level^87,88^. To account for residual dependence across experiments, studies with overlapping patient cases were assigned to a common cluster and therefore treated as a single independent unit^15,29,38^. We determined heterogeneity using a multilevel *I*^2^ ^89^. To identify sources of heterogeneity, we conducted robust meta- regressions for each categorical moderator.

All analyses were conducted in R (version 4.5.0)^90^, using metafor (version 4.8-0) for multilevel meta-analytic models^91^ and clubSandwich (version 0.7.0) for CR2 cluster-robust variance estimation with small-sample corrections^92^.

## Data Availability

All code and datasets of our meta-analysis are accessible via our GitHub repository: https://github.com/isatow/Metaanalysis_LLMs_in_Medicine

## Author Contributions

I.T. and E.E. screened studies, extracted, and analyzed the data. S.F. resolved any disagreements among I.T. and E.E.; I.T. and E.E. drafted the current manuscript, while M.S. was involved in the first draft of the manuscript. S.F., T.T., and I.M.W. edited and revised the manuscript. All authors had access to the data in the study.

## Online Appendix

### A Clinical decision support systems vs. LLMs

**Table S1.** Conceptual distinction between traditional clinical decision support systems and large language models.

| Category | Definition | Implication for the present review |
| --- | --- | --- |
| **Traditional rule-based or statistical clinical decision support systems** | Computerized systems that generate patient-specific recommendations by applying predefined logic to structured inputs, either knowledge-based rules and curated knowledge bases, or non-knowledge-based statistical and conventional (non-generative) machine-learning models; typically, they do not produce free-text natural-language output.<br>**Exemplary medical use cases:** drug–drug interaction and allergy alerts, guideline reminders<sup>1</sup> | **Excluded** unless the system embeds an LLM component whose generated output is presented to the physician. |
| **General-purpose LLMs** | LLMs pretrained via self-supervision on broad, predominantly non-domain-specific corpora that respond to free-text queries without task-specific training and generate natural-language output; in clinical use, they are prompted with patient information to support diagnostic reasoning.<br>**Exemplary medical use cases:** differential diagnosis, reasoning support, medical report summaries<sup>2</sup> | **Included** when the model’s output is provided to physicians as an assistive input to diagnosis, i.e., under physician supervision, not stand-alone. |
| **Domain-adapted or fine-tuned (medical) LLMs** | LLMs specialized for medicine through fine-tuning, instruction/prompt tuning, domain-specific pretraining, or retrieval augmentation, as well as purpose-built medical foundation models (e.g., Med-PaLM).<br>**Exemplary medical use cases:** the same diagnostic tasks as general-purpose LLMs, extended by domain-specific knowledge<sup>3</sup> | **Included** on the same basis as general-purpose LLMs. |

### B Search string

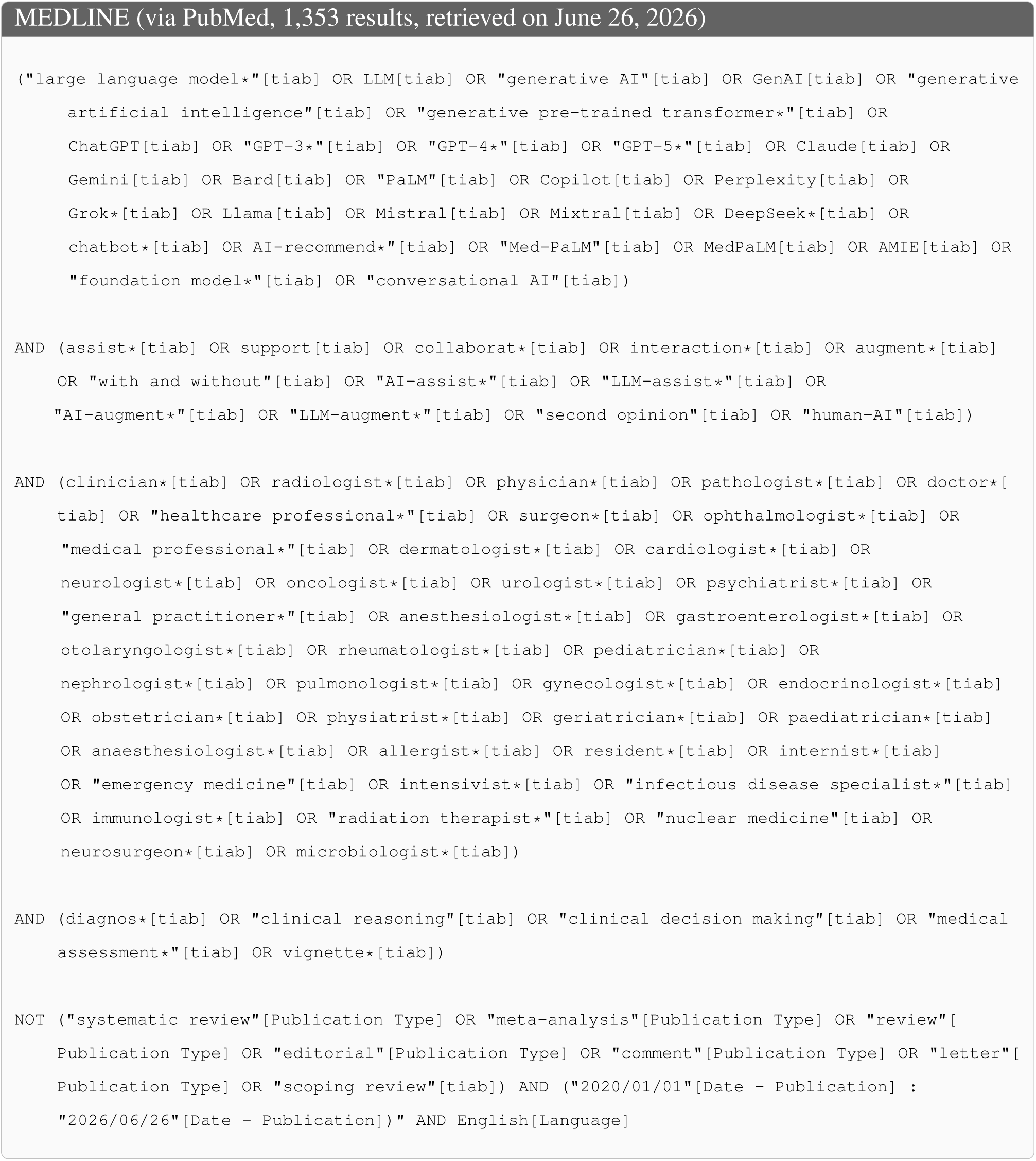
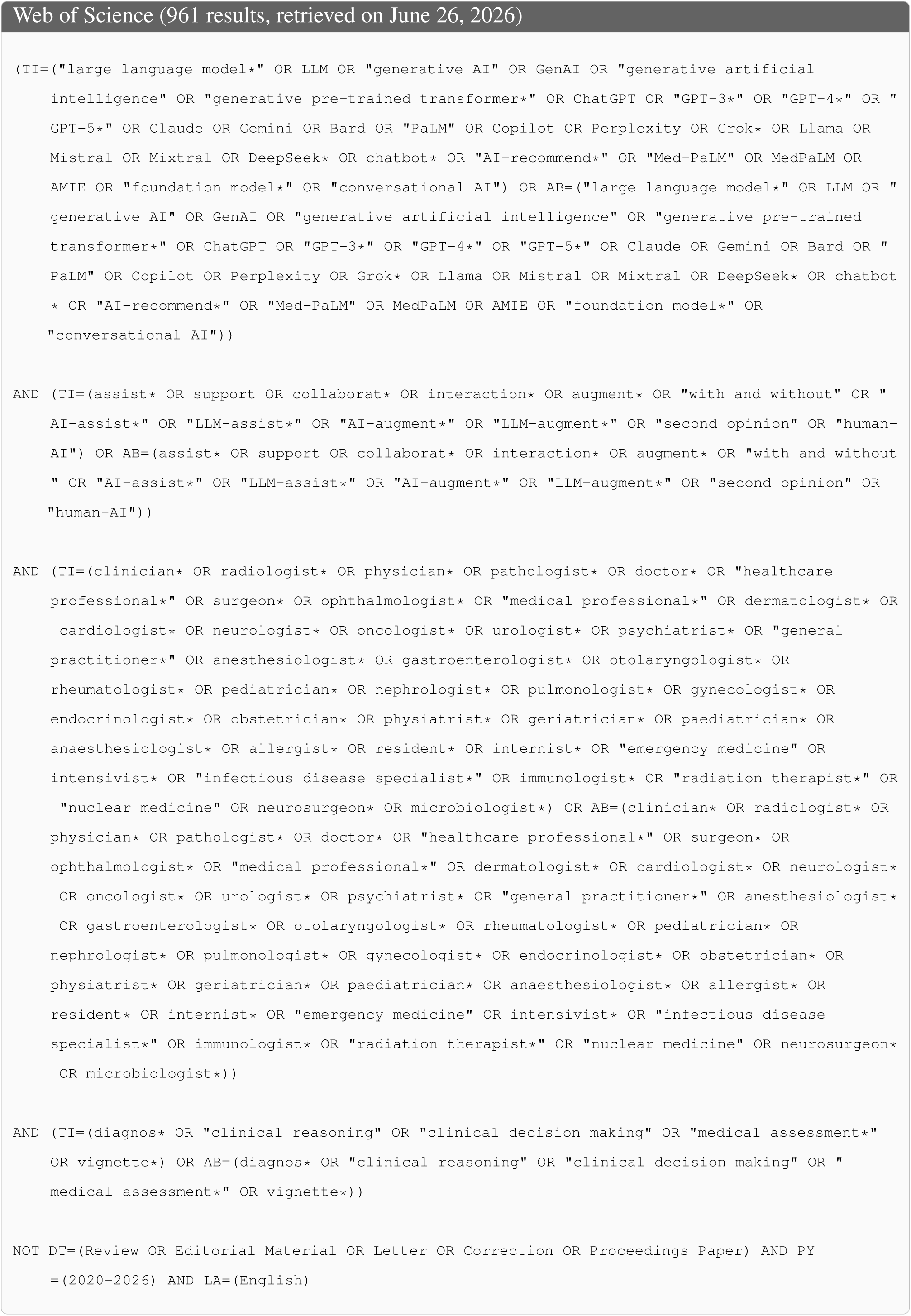
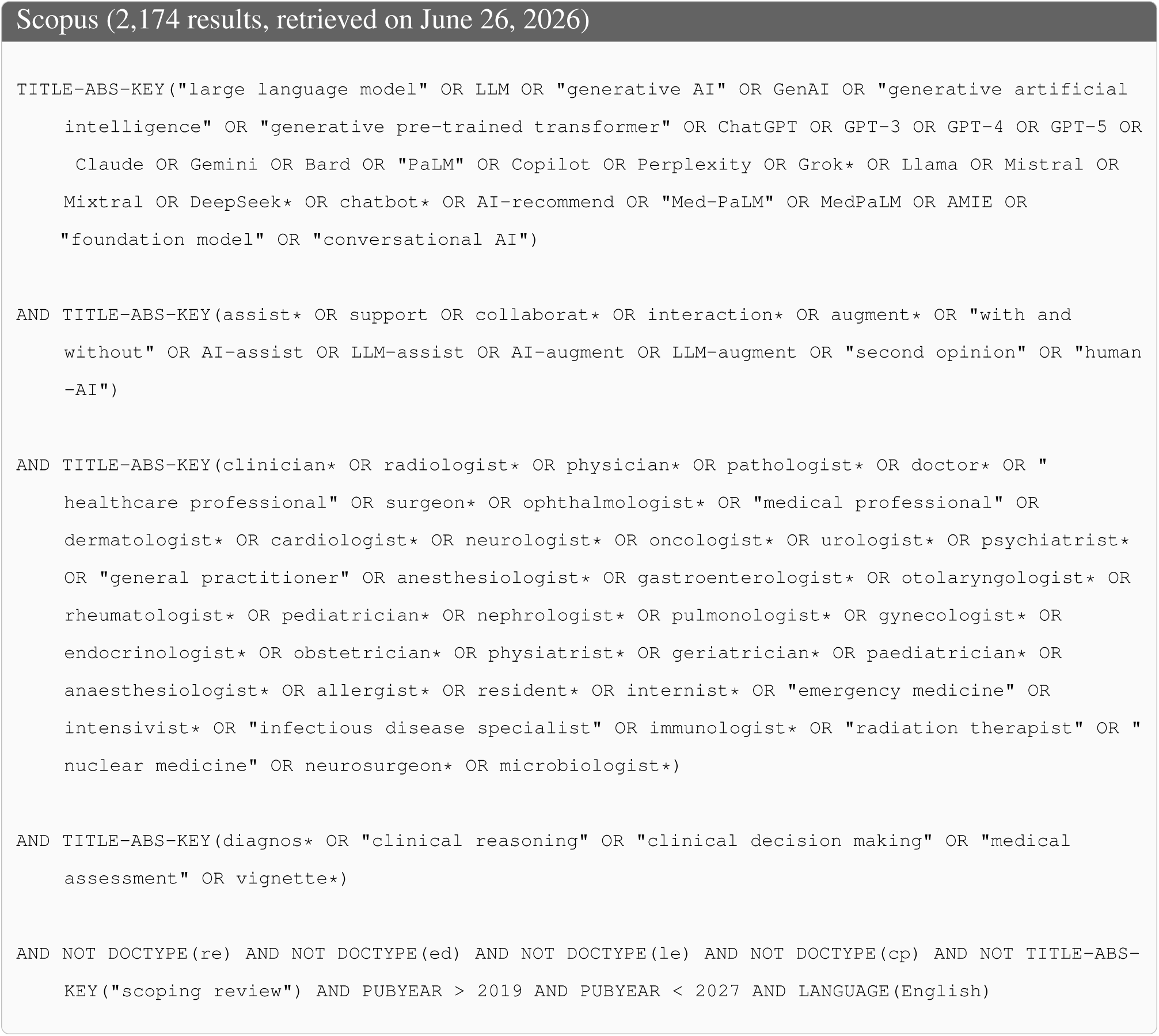
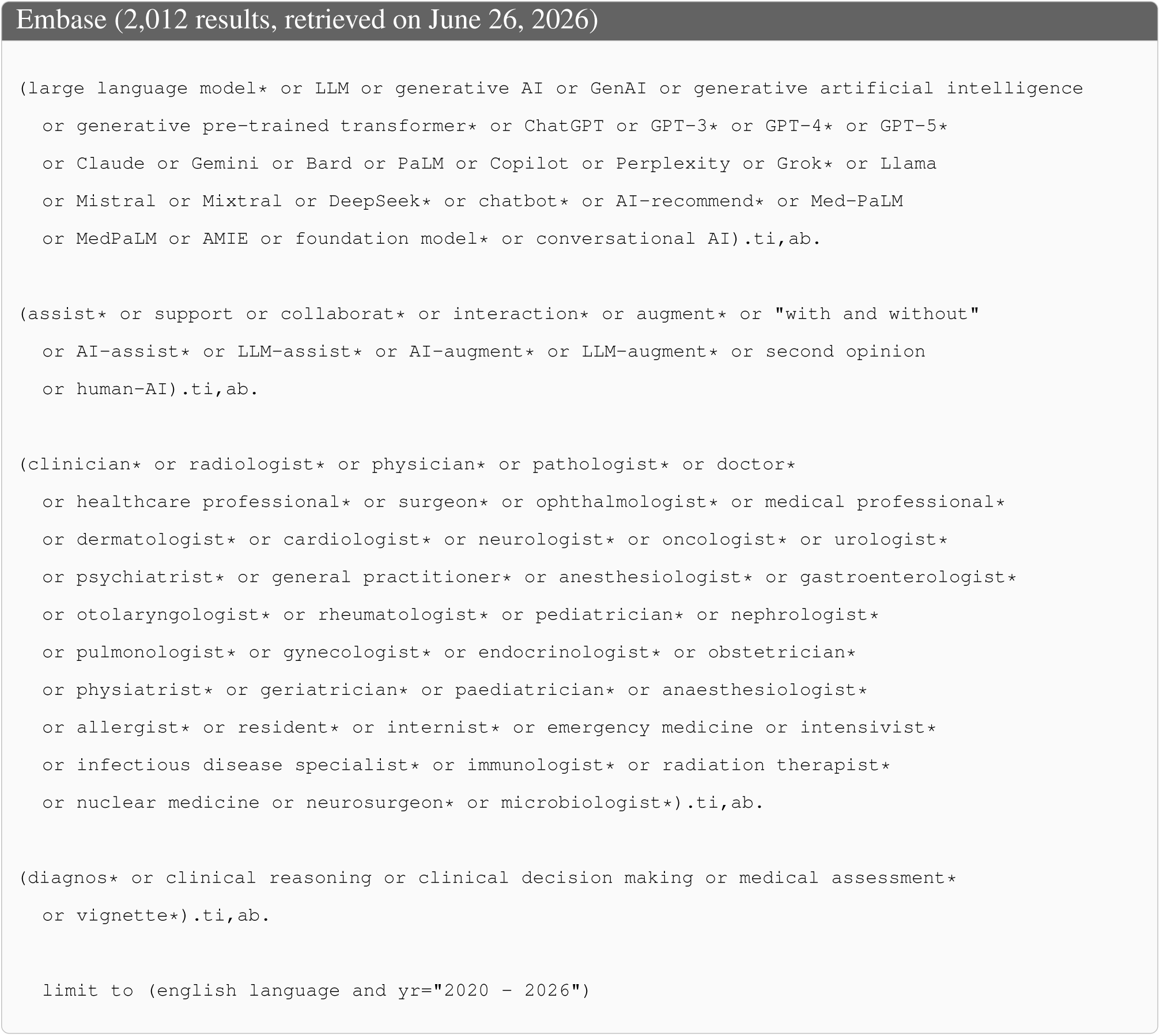
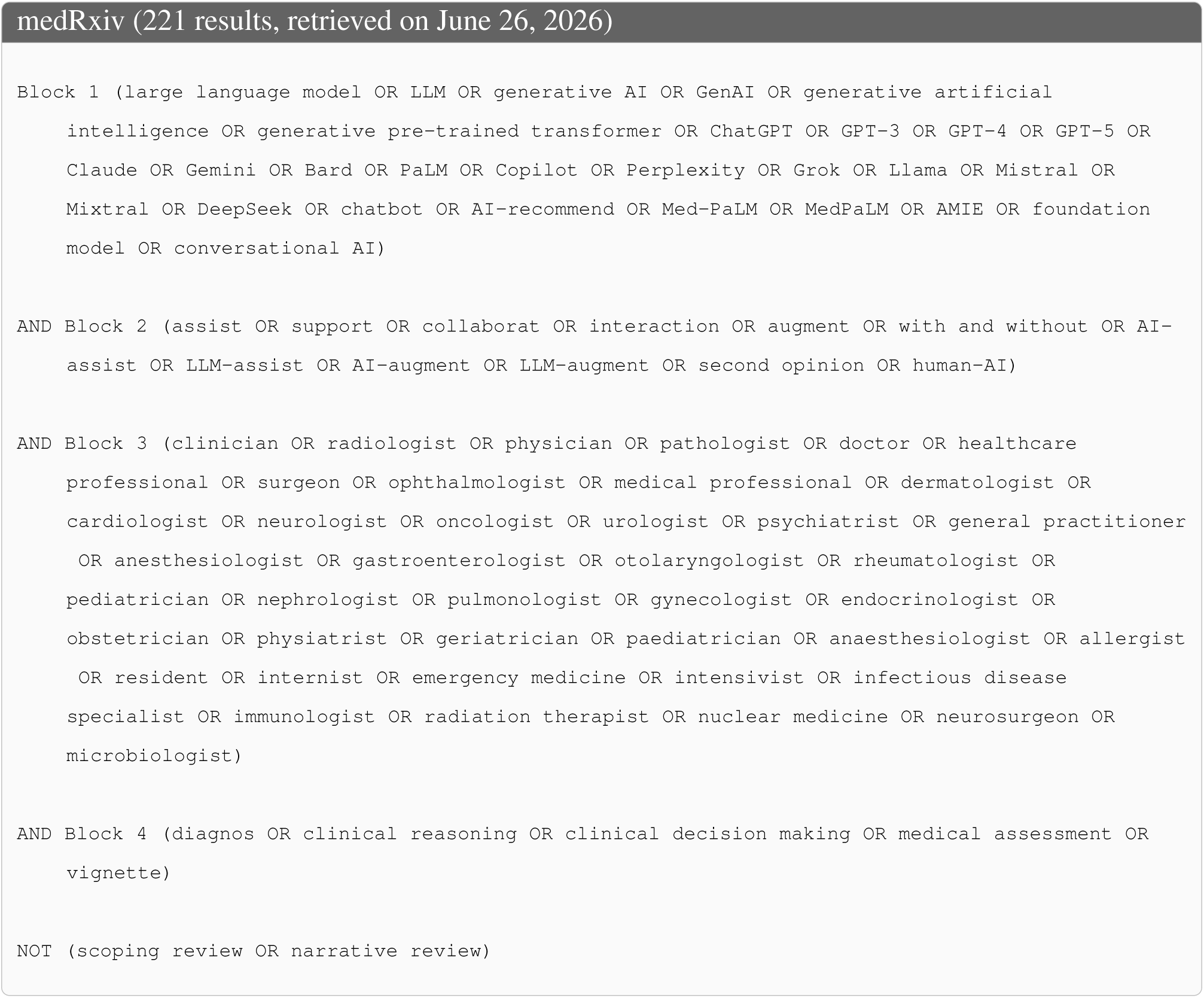

For MEDLINE, Web of Science, Scopus, and medRxiv, database searches were executed in R using the respective APIs (NCBI E-utilities, Clarivate Web of Science Starter API, Elsevier Scopus Search API, and medRxiv REST API). For MEDLINE, Web of Science, and Scopus, the search strategy was implemented as fielded Boolean queries restricted to titles and abstracts, with database-specific syntax adaptations (e.g., due to available search fields in each database). Because the medRxiv API does not support fielded Boolean queries, the search was implemented in two stages: all medRxiv preprints within the predefined date range (January 1, 2020 until June, 26, 2026) were first retrieved via the API; the retrieved records were then filtered locally in R against the same conceptual search blocks used for the other databases. Singular/stem terms were used, with plurals and other morphological variants captured automatically through case-insensitive substring matching, mirroring the truncation wildcards (e.g., `clinician`, diagnos_*_) used in the fielded-database queries. Embase was searched via the Ovid interface using the search strategy shown above (different search strategy required due to licensing restrictions).

### C Additional details on selected coding dimensions

The effect of an LLM might also depend on the **career stage**. A recent systematic review found no significant performance gap between AI and less experienced physicians (e.g., residents), but showed that AI underperformed relative to more experienced clinicians (e.g., attendings)^4^. We define physicians with less than five years of experience as residents and physicians with five or more years of experience as attendings^5^.

**Response format** differentiates the different types of LLM-generated responses. Differential diagnoses can increase the comprehensiveness of differential lists^6^, stepwise explanations (i.e., Chain-of-Thought (CoT)) improve transparency, facilitate error identification and, thus, may improve diagnostic accuracy^7^. Free text answers may provide general (pathological) information or image descriptions^5^.

The **task format** refers to the type of diagnostic question posed. We distinguish three formats: open-ended questions, e.g., asking for a differential and the most likely diagnosis^8^, multiple-choice questions which offer a predefined set of answers^9^, or classification questions, in which one of several possible categories needs to be selected (e.g., classifying a tumor as benign/malignant)^10^. In addition, the **input form of patient information** may impact LLM performance. We distinguish between purely text-based, image-based, and text- and image-based cases, given recent evidence that GPT-4V might have limitations in reliably interpreting (radiologic) images^9^.

Diagnostic accuracy is often assessed using different **accuracy measures**. Some studies evaluate whether the top-ranked diagnosis matches the gold standard (top-1 accuracy)^8^, others measure whether the correct diagnosis appeared within a differential list (e.g., top-3 accuracy)^11^. Distinguishing these accuracy measures is important as diagnostic accuracy may vary depending on the accuracy measure used. For example, accuracy tends to increase as the differential diagnosis list expands^6^.

### D Statistical analysis

Effect sizes were calculated as Hedges’ *g*, with positive values indicating higher diagnostic accuracy with LLM assistance. Calculations were performed using `metafor` and were adapted to the outcome type and study design.

For binary diagnostic accuracy outcomes, accuracy was represented as a Bernoulli outcome on the 0–1 scale. Reported percentages were converted to proportions, and the corresponding raw-score standard deviation was calculated as

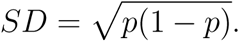

For continuous accuracy outcomes, reported standard deviations were used.

For parallel-group comparisons, we used the heteroscedastic standardized mean difference (`SMDH`) for binary outcomes and the conventional standardized mean difference (`SMD`) for continuous outcomes. For within-subject comparisons in which the same unit of analysis (case, physician, or physician–case) was assessed with and without LLM assistance, we used the paired standardized mean-change estimator implemented as `SMCRPH` in `metafor`^12^. This estimator standardizes the mean difference using the average variance of the two conditions and incorporates the within-pair correlation *r* into the sampling variance. This within-pair correlation *r* was derived from the discordant cells of the paired 2 × 2 table for binary outcomes. Otherwise, a correlation reported by the study was used. Where *r* remained unavailable, it was imputed as the median of empirical correlations for the corresponding unit of analysis, with donor values first aggregated within paper/case-corpus clusters to avoid disproportionate weighting of studies contributing multiple estimates. The resulting median correlations were *r* = 0.78 for case-level estimates, *r* = 0.94 for physician-level estimates, and *r* = 0.63 for physician–case-level estimates. Robustness to the correlation assumption was assessed by repeating the meta-analysis with *r* = 0, 0.3, 0.5, 0.7, and 0.9. The small-sample correction yielding Hedges’ *g* was applied directly by `metafor`.^13^

Several studies reported multiple case evaluations provided by the same physicians (referred to as physician–case). For these effect sizes, we multiplied the sampling variance with the design effect

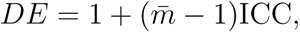

where *m̂* denotes the average number of observations per physician and ICC the intraclass correlation coefficient. Thereby, we ensure that physician–case observations are not treated as independent. Study-specific ICCs were used where available; otherwise, an ICC of 0.027^14^ was used. Robustness to this assumption was assessed by repeating the analysis with ICC = 0, 0.027, 0.05, 0.10, and 0.20.

For selected studies reporting multiple granular estimates for the same comparison, raw arm-level information was pooled before effect size calculation. Pooling accounted for the dependence structure of the source data, distinguishing, for example, multiple physicians evaluating the same cases, separate groups of physicians evaluating different cases, and different case subsets evaluated by the same physicians. Pooling was only performed when the comparison had the same values for all moderators included in the meta-regression analyses. Previously calculated effect sizes were not averaged.

Moderator analyses were conducted using separate no-intercept multilevel meta-regressions with CR2-robust inference to estimate the mean effect for each subgroup. Differences among moderator categories were assessed separately using CR2-robust Hotelling-Zhang small-sample omnibus tests^15^. For omnibus tests, sparse moderator categories represented by fewer than two independent paper/case-corpus clusters were combined into an “Other” category. As a supplementary analysis, we additionally fitted a multivariable meta-regression including all moderators simultaneously using effect coding. Moderator analyses were considered exploratory.

Sensitivity and robustness analyses restricted the analysis to experiments with low risk of bias, added a paper/case-corpus level random intercept to the primary model, varied the assumed within-subject correlation (*r*) and ICC as described above, estimated pooled effects separately by publication status (peer-reviewed vs. preprint) and study design (parallel-group vs. within-subject), and sequentially excluded individual effect sizes, experiments, and studies (leave-one-out analyses). Small-study effects were assessed using a funnel plot and a multilevel, CR2-robust Egger-type regression with 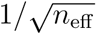 as a sample-size-based predictor, where *n*_eff_ is the number of pairs (within-subject) or the total number of observations across both arms (parallel-group), adjusted for repeated case evaluations by the same physician using the design effect described above^16^. Funnel asymmetry is quantified by the regression slope (*β*).

### E Risk-of-bias assessment (PROBAST)

**Fig. S1.**
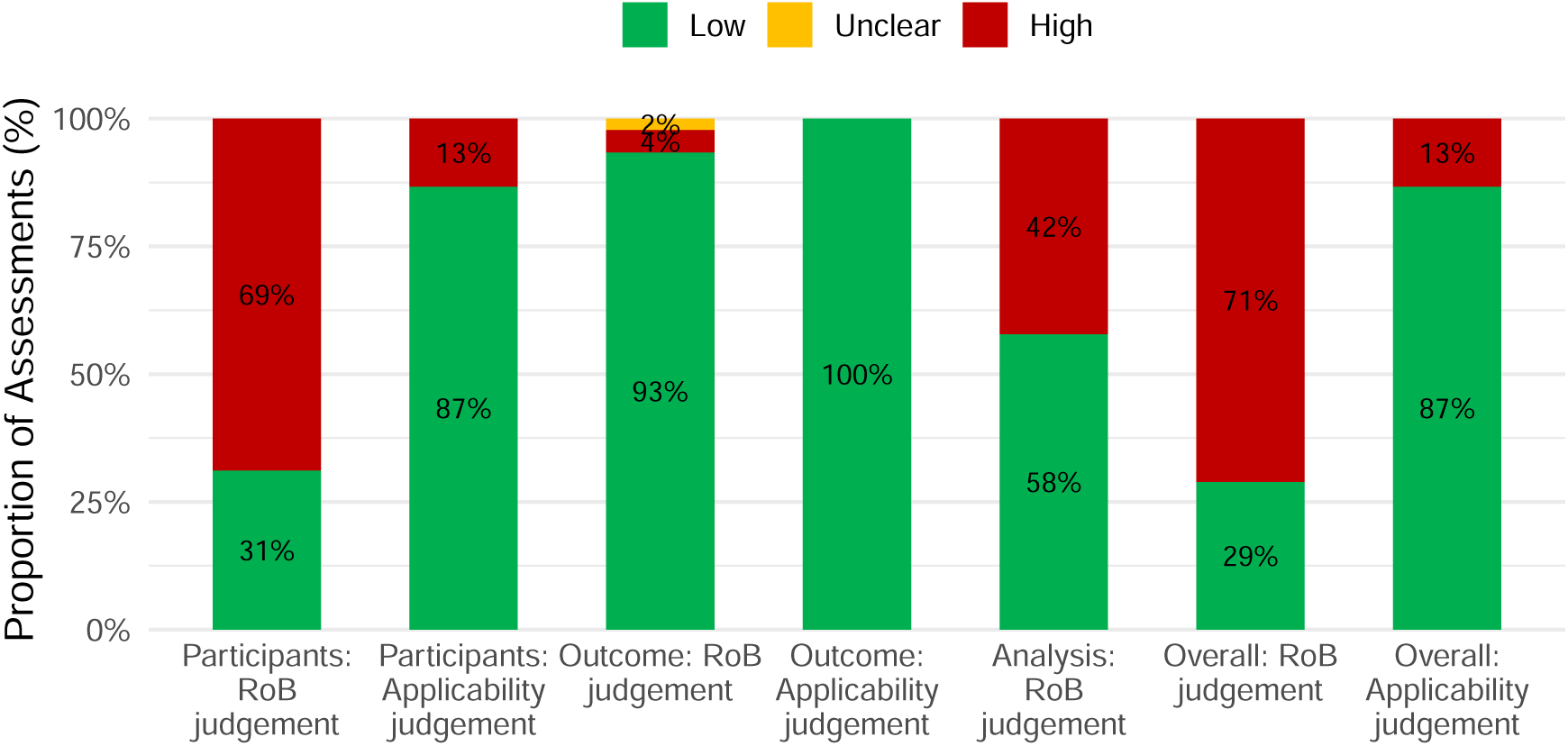
Summary of PROBAST assessment. The figure illustrates the result of our PROBAST assessment for the 42 studies included in our meta-analysis. For two articles reporting multiple distinct experiments, experiments were assessed separately because their risk-of-bias profiles differed. Hence, we conducted 45 PROBAST assessments in total. We excluded selected PROBAST questions as they are not applicable to our analysis. This involves all three questions from Domain 2 (predictors), as well as questions 3.3, 3.5, and 3.6 from Domain 3 (outcome) and questions 4.5 and 4.9 from Domain 4 (analysis)^4^.

**Fig. S2.**
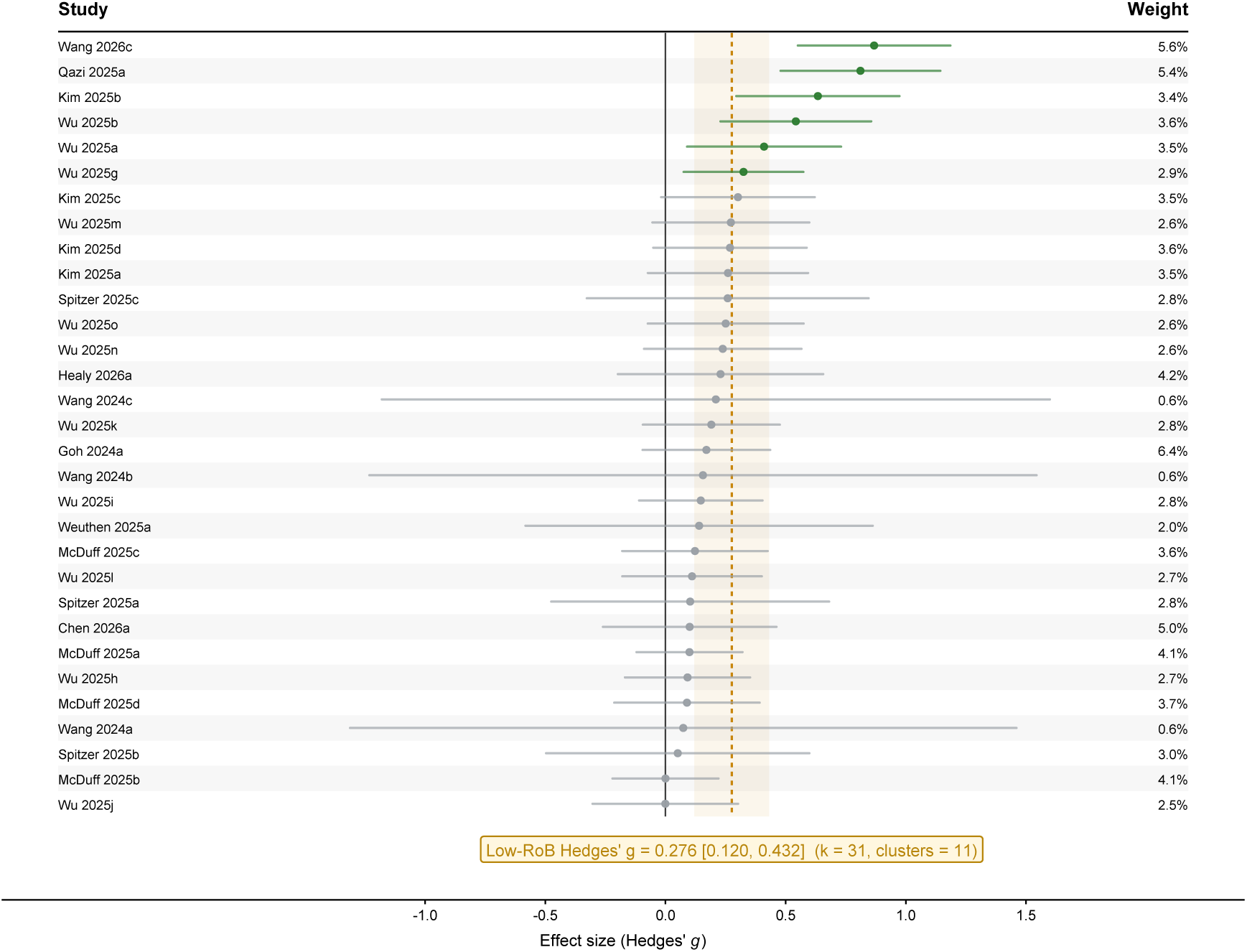
Low risk-of-bias forest plot. The forest plot includes all effect sizes from experiments judged to be at low risk-of-bias (*k* = 31 effect sizes from *n* = 12 studies, grouped into 11 paper/case-corpus clusters as two studies drawing on the same, partly overlapping case corpus were combined into a single cluster to account for cross-study dependence^17,18^). Points indicate individual effect sizes (Hedges’ *g*) and horizontal lines their 95% CIs. Green points indicate effect sizes with 95% CIs above zero, while grey points indicate CIs including zero. The solid vertical line marks *g* = 0. The dashed yellow line and shaded band indicate the pooled effect and its 95% CI (Hedges’ *g* = 0.28, 95% CI [0.12, 0.43]). Labels on the left refer to the study from which the effect size was derived; percentages on the right indicate the weight of the respective effect size.

### F Supplementary analyses

**Fig. S3.**
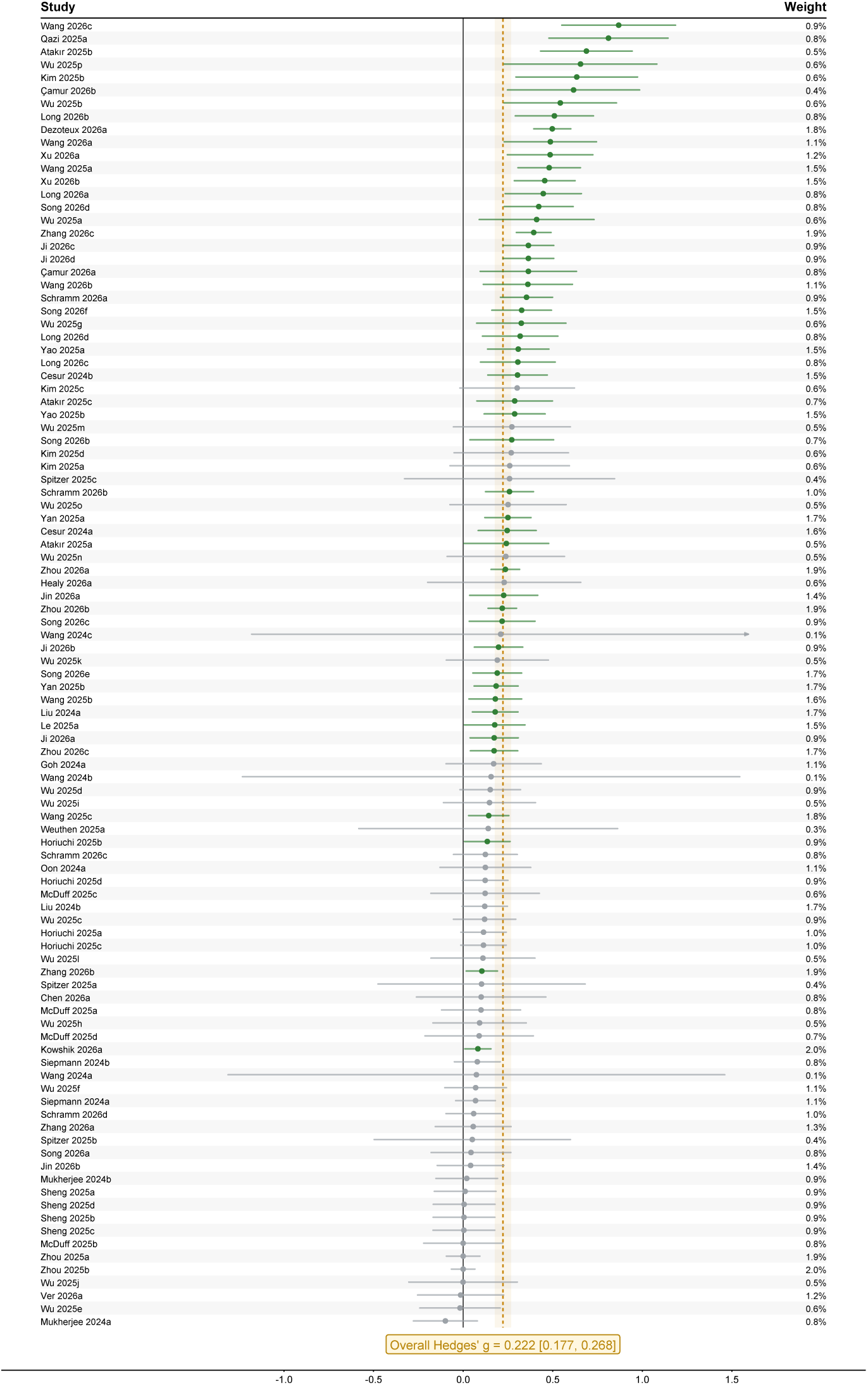
Forest plot of effect sizes (*k* = 101) comparing diagnostic accuracy with and without LLM assistance (including study and weight labels). Each horizontal line represents the 95% CI for an individual effect size, with the central marker indicating Hedges’ *g*. The pooled effect size (Hedges’ *g* = 0.22, 95% CI [0.18, 0.27]) is indicated by the dashed vertical line and yellow-shaded confidence band. The gray line corresponds to an effect size of *g* = 0. Labels on the left-hand side refer to the study from which the effect size was derived; percentages on the right indicate the weight of the respective effect size.

**Table S2.** Omnibus tests of differences between moderator subgroups.

| Moderator | Groups | $F(df_1, df_2)$ | $p$ |
| --- | --- | --- | --- |
| **Accuracy measure** | 3 | $F(2, 1.59) = 0.358$ | 0.744 |
| **LLM model** | 5 | $F(4, 1.67) = 1.991$ | 0.391 |
| **Task format** | 3 | $F(2, 12.21) = 0.505$ | 0.616 |
| **Input form** | 3 | $F(2, 12.12) = 0.046$ | 0.955 |
| **Response format** | 4 | $F(3, 13.83) = 1.552$ | 0.246 |
| **Customized prompt** | 2 | $F(1, 12.70) = 0.645$ | 0.437 |
| **Medical field** | 4 | $F(3, 3.83) = 0.715$ | 0.594 |
| **Career stage** | 3 | $F(2, 9.84) = 3.084$ | 0.091 |
| **Setting of patient cases** | 3 | $F(2, 6.53) = 0.093$ | 0.913 |

*Note.* Analyses are based on 42 studies with 101 effect sizes and pool across diagnostic accuracy measures unless the accuracy measure itself is examined. Omnibus tests assess whether mean effect sizes differ across moderator subgroups using CR2-robust Hotelling-Zhang (HTZ) inference clustered at the paper/case-corpus level. Groups denotes the number of moderator subgroups included in the respective test. $df_1$ denotes the degrees of freedom and equals Groups $-1$ . $df_2$ denotes the HTZ-adjusted degrees of freedom and reflects the effective amount of independent information available for the subgroup comparison after accounting for clustering and the small-sample CR2 adjustment; it may therefore be non-integer. $F$ quantifies the magnitude of the differences between subgroup means relative to their uncertainty. $p$ is the corresponding omnibus $p$ -value and tests the null hypothesis that the moderator subgroup means do not differ. Sparse moderator categories (i.e., represented by fewer than two independent paper/case-corpus clusters) are collapsed.

**Table S3.** Meta-regression results across all moderators.

| Moderator | Subgroup | Deviation from mean ($\beta$) | SE | 95% CI |
| --- | --- | --- | --- | --- |
| Accuracy measure | Other | 0.008 | 0.012 | [-0.09, 0.11] |
| Accuracy measure | Top-1 | -0.028 | 0.013 | [-0.10, 0.04] |
| Accuracy measure | Top-3 | 0.020 | 0.019 | [-0.04, 0.08] |
| LLM model | DeepSeek-R1 | 0.249 | 0.100 | [-0.41, 0.91] |
| LLM model | GPT-4 | -0.153 | 0.082 | [-0.34, 0.04] |
| LLM model | GPT-4o | 0.111 | 0.081 | [-0.07, 0.29] |
| LLM model | GPT-4V | -0.226 | 0.119 | [-0.69, 0.24] |
| LLM model | Other | 0.019 | 0.057 | [-0.12, 0.15] |
| Task format | Classification | 0.005 | 0.055 | [-0.13, 0.14] |
| Task format | Multiple choice | 0.020 | 0.036 | [-0.07, 0.11] |
| Task format | Open-ended | -0.025 | 0.055 | [-0.15, 0.10] |
| Input form | Image | 0.018 | 0.063 | [-0.13, 0.16] |
| Input form | Multimodal (text + image) | 0.006 | 0.045 | [-0.09, 0.10] |
| Input form | Text | -0.024 | 0.060 | [-0.15, 0.11] |
| Response format | CoT | 0.105 | 0.066 | [-0.04, 0.25] |
| Response format | Differential | -0.021 | 0.052 | [-0.13, 0.09] |
| Response format | Differential, free text | -0.103 | 0.049 | [-0.21, 0.01] |
| Response format | Free text | 0.018 | 0.061 | [-0.12, 0.15] |
| Customized prompt | No | -0.063 | 0.038 | [-0.15, 0.02] |
| Customized prompt | Yes | 0.063 | 0.038 | [-0.02, 0.15] |
| Medical field | General medicine | 0.026 | 0.097 | [-0.20, 0.25] |
| Medical field | Ophthalmology | 0.026 | 0.090 | [-0.19, 0.24] |
| Medical field | Other | -0.075 | 0.073 | [-0.23, 0.08] |
| Medical field | Radiology | 0.022 | 0.054 | [-0.10, 0.14] |
| Career stage | Attending | -0.071 | 0.040 | [-0.16, 0.01] |
| Career stage | Mixed | 0.037 | 0.076 | [-0.13, 0.20] |
| Career stage | Resident | 0.034 | 0.044 | [-0.06, 0.13] |
| Setting of patient cases | Adapted real-world cases | 0.037 | 0.088 | [-0.16, 0.23] |
| Setting of patient cases | Real-world cases | -0.065 | 0.059 | [-0.19, 0.06] |
| Setting of patient cases | Published cases | 0.028 | 0.047 | [-0.08, 0.13] |
| Intercept | Grand mean | 0.311** | 0.074 | [0.15, 0.47] |

*Note.* The exploratory meta-regression is based on 42 studies with 101 effect sizes and pools across diagnostic accuracy measures. All moderators were entered simultaneously into the model. Sparse moderator categories were collapsed into an “Other” category, coefficients for “Other” therefore aggregate heterogeneous categories. Categorical moderators were effect-coded (i.e., the grand mean represents the unweighted mean across moderators); coefficients represent deviations from this mean. Standard errors shown are CR2-robust standard errors (clustered at the paper/case-corpus level, i.e., paper level except for studies sharing the same case corpus).
Significance levels: \* $p < 0.05$ , \*\* $p < 0.01$ , \*\*\* $p < 0.001$ .

### G Results of robustness and sensitivity checks

#### G.1 Robustness checks

**Fig. S4.**
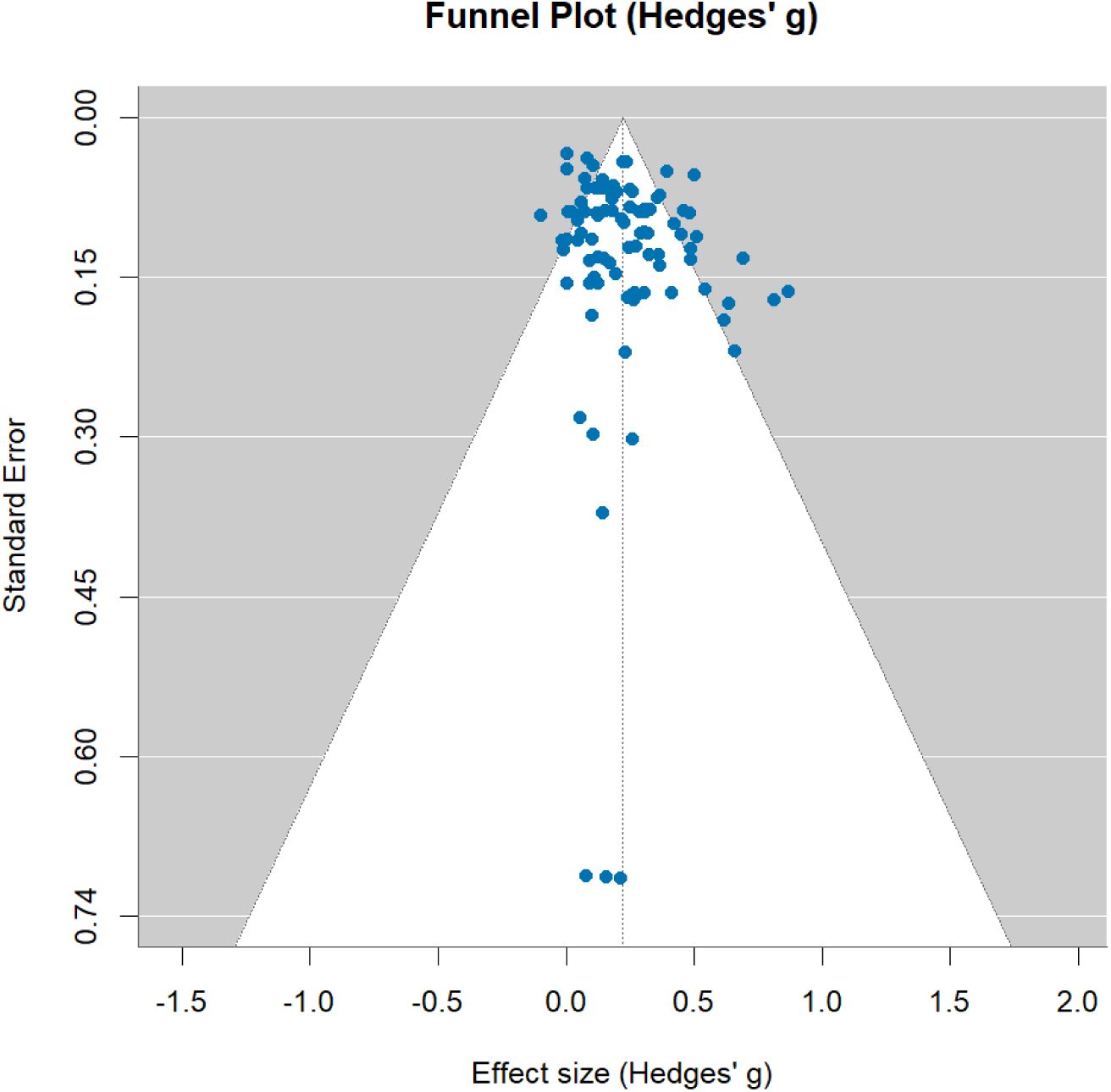
Funnel plot of effect sizes. This plot is used to assess small-study effects across the *k* = 101 effect sizes. We used the same three-level meta-analytic model as for our main analysis. We did not find evidence of substantial asymmetry.

#### G.2 Sensitivity checks

**Table S4.** Sensitivity analyses for assumptions about within-pair correlation and within-physician clustering.

| Assumed value | $k$ | Adjusted effect sizes | Hedges' $g$ | 95% CI | $\tau^2$ |
| --- | --- | --- | --- | --- | --- |
| **A. Imputed within-pair correlation $r$** |  |  |  |  |  |
| 0.0 | 101 | — | 0.222 | [0.175, 0.270] | 0.018 |
| 0.3 | 101 | — | 0.222 | [0.175, 0.269] | 0.019 |
| 0.5 | 101 | — | 0.223 | [0.176, 0.269] | 0.019 |
| 0.7 | 101 | — | 0.223 | [0.176, 0.270] | 0.020 |
| 0.9 | 101 | — | 0.223 | [0.176, 0.269] | 0.021 |
| **B. ICC / design-effect correction** |  |  |  |  |  |
| 0 | 101 | 4 | 0.229 | [0.181, 0.276] | 0.025 |
| 0.027 | 101 | 69 | 0.222 | [0.177, 0.268] | 0.019 |
| 0.05 | 101 | 69 | 0.218 | [0.173, 0.263] | 0.017 |
| 0.1 | 101 | 69 | 0.212 | [0.167, 0.258] | 0.014 |
| 0.2 | 101 | 69 | 0.207 | [0.159, 0.256] | 0.012 |
| 0 (all rows) | 101 | 0 | 0.231 | [0.183, 0.279] | 0.027 |

*Note.* Panel A varies the assumed within-pair correlation $r$ only for effect sizes for which $r$ had to be imputed; reported correlations and correlations derived from discordant paired observations remain unchanged. Panel B varies the ICC used to account for the dependency arising when the same physician contributed multiple case assessments (when no study-specific ICC was available). Study-specific ICCs remain unchanged, except in the last scenario, in which the correction is removed from all effect sizes. $k$ denotes the number of effect sizes; Hedges' $g$ the pooled effect estimate; 95% CI the corresponding confidence interval; and $\tau^2$ the total estimated heterogeneity.

**Table S5.** Sensitivity analyses by publication status and study design.

| Subgroup | $k$ | Experiments | Clusters | Hedges' $g$ | 95% CI |
| --- | --- | --- | --- | --- | --- |
| **A. Publication status** |  |  |  |  |  |
| Peer-reviewed | 97 | 67 | 37 | 0.227 | [0.177, 0.276] |
| Preprint | 4 | 4 | 3 | 0.176 | [-0.093, 0.446] |
| **B. Study design** |  |  |  |  |  |
| Parallel-group | 27 | 18 | 10 | 0.285 | [0.089, 0.480] |
| Within-subject | 74 | 53 | 31 | 0.213 | [0.164, 0.262] |

*Note.* Each row reports a separate subgroup-specific three-level meta-analytic estimate with CR2-robust inference clustered at the paper/case-corpus level. $k$ denotes the number of effect sizes; Experiments denotes the number of distinct experiments; and Clusters denotes the number of paper/shared-case-corpus clusters contributing to the respective estimate. The peer-reviewed row represents the sensitivity analysis excluding preprints. Parallel-group and within-subject rows show study design-specific pooled estimates. These estimates are descriptive subgroup analyses and do not by themselves test whether publication-status or study-design subgroups differ statistically.

**Fig. S5.**
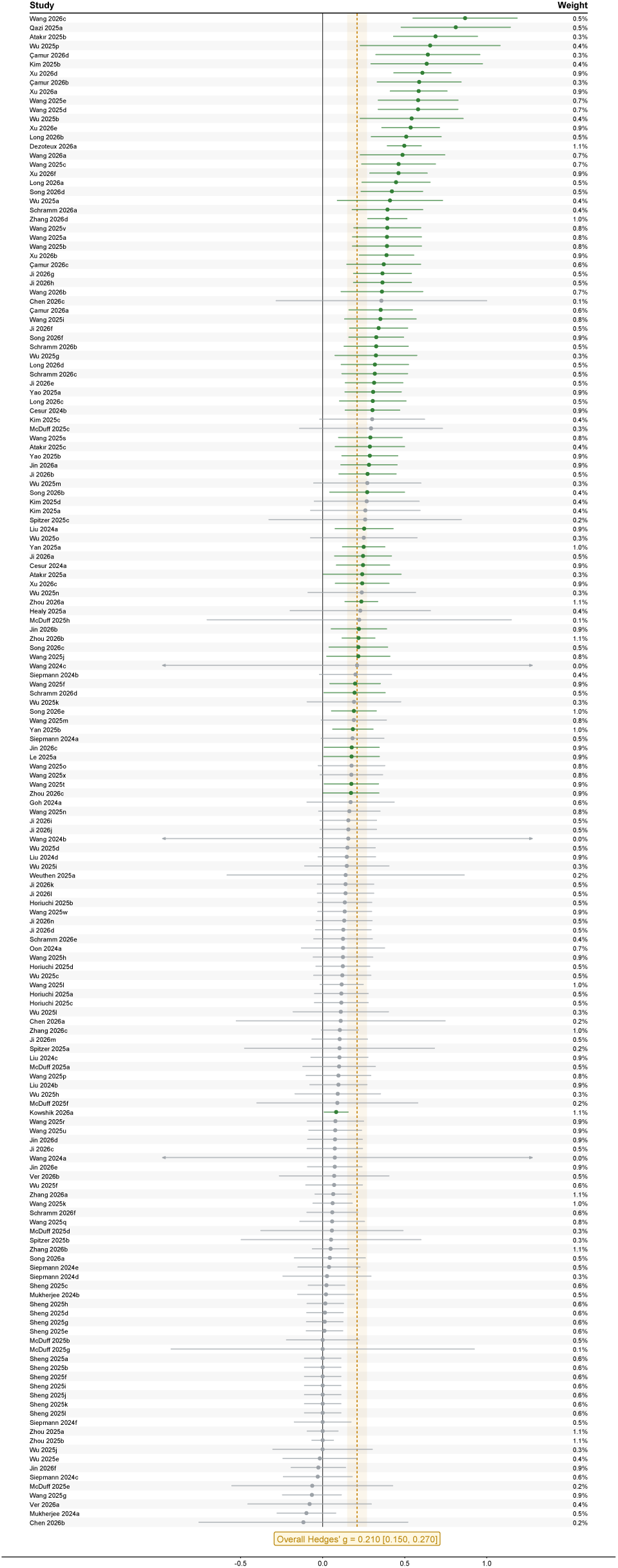
Forest plot of effect sizes (*k* = 166) comparing diagnostic accuracy with and without LLM assistance, coding each study at highest reported granularity level. Each horizontal line represents the 95% CI for an individual effect size, with the central marker indicating Hedges’ *g*. The pooled effect size (Hedges’ *g* = 0.21, 95% CI [0.15, 0.27]) is indicated by the dashed vertical line and yellow-shaded confidence band. The gray line corresponds to an effect size of *g* = 0. Labels on the left-hand side refer to the study from which the effect size was derived; labels on the right indicate the weight of the respective effect size. The figure shows all effect sizes at the most granular level reported in each study (e.g., per-physician accuracies, where reported).

### Leave-one-out Analysis

We conducted three leave-one-out analyses to assess the robustness of our results: leaving out an effect size (Supplementary Table S6), an experiment (Supplementary Table S7), and a paper (Supplementary Table S8). For each omission, we re-fitted the three-level model and re-computed the CR2-adjusted pooled *g*, the 95% CI, and total *I*^2^. The first column identifies the omitted effect size (ES), experiment, or study, followed by the number of effect sizes included in each iteration (*k*), and paper/case-corpus clusters. *SE* denotes the CR2-adjusted standard error, *t* the corresponding test statistic, and *df* the Satterthwaite degrees of freedom used for the *t*-based confidence intervals and *p*-values.

**Table S6.** Leave-one-out results: Effect size.

| Left-out<br>ES ID | $k$ | Clusters | Hedges' $g$ | $SE$ | $t$ | $df$ | $p$ | 95% CI | $I^2$ (%) |
| --- | --- | --- | --- | --- | --- | --- | --- | --- | --- |
| 5 | 100 | 40 | 0.222 | 0.022 | 9.966 | 26.00 | < 0.001 | [0.177, 0.268] | 73.4 |
| 6 | 100 | 40 | 0.221 | 0.022 | 10.013 | 26.09 | < 0.001 | [0.175, 0.266] | 72.8 |
| 11 | 100 | 39 | 0.223 | 0.022 | 9.982 | 25.84 | < 0.001 | [0.177, 0.269] | 73.5 |
| 25 | 100 | 40 | 0.223 | 0.022 | 9.998 | 26.20 | < 0.001 | [0.177, 0.268] | 73.1 |
| 26 | 100 | 40 | 0.222 | 0.022 | 10.002 | 26.21 | < 0.001 | [0.177, 0.268] | 73.1 |
| 39 | 100 | 40 | 0.223 | 0.023 | 9.874 | 25.91 | < 0.001 | [0.176, 0.269] | 73.6 |
| 42 | 100 | 40 | 0.225 | 0.022 | 10.027 | 25.88 | < 0.001 | [0.179, 0.271] | 73.2 |
| 51 | 100 | 40 | 0.223 | 0.023 | 9.907 | 25.96 | < 0.001 | [0.177, 0.270] | 73.5 |
| 52 | 100 | 40 | 0.224 | 0.023 | 9.966 | 25.95 | < 0.001 | [0.178, 0.271] | 73.3 |
| 61 | 100 | 40 | 0.223 | 0.022 | 10.015 | 26.16 | < 0.001 | [0.177, 0.268] | 73.3 |
| 62 | 100 | 40 | 0.223 | 0.022 | 10.050 | 26.15 | < 0.001 | [0.177, 0.269] | 73.3 |
| 71 | 100 | 40 | 0.221 | 0.022 | 10.013 | 26.25 | < 0.001 | [0.176, 0.267] | 73.0 |
| 74 | 100 | 40 | 0.223 | 0.022 | 9.981 | 26.30 | < 0.001 | [0.177, 0.269] | 73.3 |
| 77 | 100 | 40 | 0.223 | 0.022 | 10.088 | 26.18 | < 0.001 | [0.177, 0.268] | 73.0 |
| 78 | 100 | 40 | 0.223 | 0.022 | 10.106 | 26.18 | < 0.001 | [0.178, 0.268] | 72.9 |
| 83 | 100 | 40 | 0.223 | 0.022 | 10.100 | 26.18 | < 0.001 | [0.178, 0.268] | 72.9 |
| 84 | 100 | 40 | 0.223 | 0.022 | 10.097 | 26.18 | < 0.001 | [0.177, 0.268] | 72.9 |
| 89 | 100 | 40 | 0.223 | 0.022 | 10.034 | 26.14 | < 0.001 | [0.177, 0.268] | 72.8 |
| 92 | 100 | 40 | 0.222 | 0.022 | 10.017 | 26.17 | < 0.001 | [0.177, 0.268] | 73.0 |
| 104 | 100 | 39 | 0.225 | 0.022 | 10.069 | 25.66 | < 0.001 | [0.179, 0.271] | 73.1 |
| 109 | 100 | 39 | 0.218 | 0.022 | 9.889 | 25.51 | < 0.001 | [0.173, 0.263] | 72.3 |
| 111 | 100 | 40 | 0.223 | 0.022 | 9.982 | 26.19 | < 0.001 | [0.177, 0.269] | 73.7 |
| 112 | 100 | 40 | 0.220 | 0.022 | 10.010 | 26.13 | < 0.001 | [0.175, 0.265] | 72.7 |
| 113 | 100 | 40 | 0.223 | 0.022 | 9.983 | 26.17 | < 0.001 | [0.177, 0.269] | 73.6 |
| 114 | 100 | 40 | 0.223 | 0.023 | 9.906 | 25.98 | < 0.001 | [0.177, 0.270] | 73.5 |
| 121 | 100 | 40 | 0.224 | 0.023 | 9.930 | 25.93 | < 0.001 | [0.178, 0.270] | 73.3 |
| 153 | 100 | 40 | 0.219 | 0.022 | 10.145 | 25.90 | < 0.001 | [0.175, 0.263] | 72.8 |
| 155 | 100 | 40 | 0.219 | 0.022 | 10.128 | 25.77 | < 0.001 | [0.174, 0.263] | 72.5 |
| 164 | 100 | 40 | 0.225 | 0.023 | 9.968 | 26.37 | < 0.001 | [0.178, 0.271] | 73.3 |
| 311 | 100 | 40 | 0.222 | 0.023 | 9.863 | 25.92 | < 0.001 | [0.176, 0.269] | 73.6 |
| 321 | 100 | 40 | 0.221 | 0.022 | 9.850 | 25.92 | < 0.001 | [0.175, 0.267] | 73.4 |
| 511 | 100 | 39 | 0.216 | 0.022 | 9.930 | 25.34 | < 0.001 | [0.172, 0.261] | 71.1 |
| 611 | 100 | 39 | 0.223 | 0.022 | 9.933 | 25.72 | < 0.001 | [0.177, 0.269] | 73.6 |

Table S6. Leave-one-out results: Effect size (*continued*)
| Left-out<br>ES ID | $k$ | Clusters | Hedges' $g$ | $SE$ | $t$ | $df$ | $p$ | 95% CI | $I^2$ (%) |
| --- | --- | --- | --- | --- | --- | --- | --- | --- | --- |
| 711 | 100 | 39 | 0.222 | 0.022 | 9.957 | 25.91 | < 0.001 | [0.177, 0.268] | 73.5 |
| 811 | 100 | 40 | 0.223 | 0.022 | 10.041 | 26.20 | < 0.001 | [0.177, 0.268] | 73.0 |
| 812 | 100 | 40 | 0.222 | 0.022 | 10.001 | 26.21 | < 0.001 | [0.177, 0.268] | 73.0 |
| 821 | 100 | 40 | 0.223 | 0.022 | 10.031 | 26.20 | < 0.001 | [0.177, 0.268] | 73.0 |
| 822 | 100 | 40 | 0.222 | 0.022 | 10.014 | 26.21 | < 0.001 | [0.177, 0.268] | 73.0 |
| 941 | 100 | 40 | 0.222 | 0.022 | 10.001 | 26.20 | < 0.001 | [0.176, 0.268] | 73.0 |
| 942 | 100 | 40 | 0.222 | 0.022 | 10.001 | 26.20 | < 0.001 | [0.176, 0.268] | 73.0 |
| 1111 | 100 | 40 | 0.223 | 0.022 | 9.964 | 26.36 | < 0.001 | [0.177, 0.269] | 73.7 |
| 1112 | 100 | 40 | 0.220 | 0.022 | 9.978 | 26.29 | < 0.001 | [0.175, 0.265] | 72.8 |
| 1211 | 100 | 40 | 0.222 | 0.022 | 9.990 | 26.33 | < 0.001 | [0.176, 0.268] | 73.3 |
| 1212 | 100 | 40 | 0.222 | 0.022 | 9.985 | 26.33 | < 0.001 | [0.177, 0.268] | 73.4 |
| 1311 | 100 | 39 | 0.225 | 0.023 | 9.989 | 25.43 | < 0.001 | [0.179, 0.272] | 72.4 |
| 1411 | 100 | 39 | 0.223 | 0.023 | 9.895 | 25.58 | < 0.001 | [0.177, 0.270] | 73.6 |
| 1611 | 100 | 40 | 0.222 | 0.022 | 10.021 | 26.16 | < 0.001 | [0.176, 0.267] | 73.0 |
| 1612 | 100 | 40 | 0.221 | 0.022 | 10.073 | 26.14 | < 0.001 | [0.176, 0.266] | 72.6 |
| 1621 | 100 | 40 | 0.222 | 0.022 | 10.004 | 26.18 | < 0.001 | [0.177, 0.268] | 73.2 |
| 1622 | 100 | 40 | 0.222 | 0.022 | 10.018 | 26.18 | < 0.001 | [0.176, 0.268] | 73.2 |
| 1711 | 100 | 40 | 0.222 | 0.022 | 9.978 | 26.17 | < 0.001 | [0.177, 0.268] | 73.3 |
| 1712 | 100 | 40 | 0.223 | 0.022 | 10.117 | 26.14 | < 0.001 | [0.178, 0.269] | 73.0 |
| 1811 | 100 | 40 | 0.224 | 0.022 | 10.160 | 26.08 | < 0.001 | [0.178, 0.269] | 72.4 |
| 1813 | 100 | 40 | 0.222 | 0.022 | 9.967 | 26.13 | < 0.001 | [0.176, 0.268] | 73.4 |
| 1911 | 100 | 39 | 0.224 | 0.022 | 9.955 | 25.70 | < 0.001 | [0.177, 0.270] | 73.5 |
| 2012 | 100 | 39 | 0.217 | 0.022 | 10.007 | 25.70 | < 0.001 | [0.172, 0.261] | 71.6 |
| 2131 | 100 | 40 | 0.222 | 0.022 | 10.004 | 26.27 | < 0.001 | [0.177, 0.268] | 73.3 |
| 2132 | 100 | 40 | 0.223 | 0.022 | 9.957 | 26.29 | < 0.001 | [0.177, 0.269] | 72.9 |
| 2411 | 100 | 40 | 0.224 | 0.022 | 9.994 | 26.18 | < 0.001 | [0.178, 0.270] | 73.3 |
| 2412 | 100 | 40 | 0.221 | 0.022 | 9.951 | 26.20 | < 0.001 | [0.176, 0.267] | 73.6 |
| 2421 | 100 | 40 | 0.224 | 0.022 | 10.006 | 26.21 | < 0.001 | [0.178, 0.270] | 73.7 |
| 2422 | 100 | 40 | 0.221 | 0.022 | 9.927 | 26.17 | < 0.001 | [0.175, 0.266] | 73.0 |
| 2511 | 100 | 40 | 0.223 | 0.023 | 9.854 | 25.91 | < 0.001 | [0.177, 0.270] | 73.5 |
| 2521 | 100 | 40 | 0.221 | 0.022 | 9.821 | 25.94 | < 0.001 | [0.175, 0.267] | 73.3 |
| 2611 | 100 | 40 | 0.223 | 0.022 | 10.004 | 26.06 | < 0.001 | [0.177, 0.269] | 73.4 |
| 2621 | 100 | 40 | 0.223 | 0.022 | 10.011 | 26.04 | < 0.001 | [0.177, 0.269] | 73.4 |
| 2631 | 100 | 40 | 0.222 | 0.022 | 9.975 | 26.06 | < 0.001 | [0.176, 0.268] | 73.4 |
| 2811 | 100 | 40 | 0.219 | 0.022 | 9.999 | 25.85 | < 0.001 | [0.174, 0.264] | 72.8 |
| 2821 | 100 | 40 | 0.221 | 0.022 | 9.978 | 25.87 | < 0.001 | [0.175, 0.266] | 73.3 |
| 2911 | 100 | 39 | 0.216 | 0.021 | 10.056 | 25.64 | < 0.001 | [0.172, 0.260] | 70.9 |
| 3211 | 100 | 40 | 0.223 | 0.022 | 10.016 | 26.16 | < 0.001 | [0.177, 0.268] | 73.3 |
| 3221 | 100 | 40 | 0.222 | 0.022 | 10.014 | 26.16 | < 0.001 | [0.177, 0.268] | 73.3 |
| 3231 | 100 | 40 | 0.222 | 0.022 | 10.012 | 26.16 | < 0.001 | [0.177, 0.268] | 73.3 |
| 3311 | 100 | 39 | 0.223 | 0.022 | 10.001 | 26.07 | < 0.001 | [0.177, 0.268] | 73.4 |
| 3411 | 100 | 40 | 0.222 | 0.022 | 9.948 | 26.48 | < 0.001 | [0.176, 0.268] | 73.2 |
| 3412 | 100 | 40 | 0.221 | 0.023 | 9.793 | 26.47 | < 0.001 | [0.174, 0.267] | 72.8 |
| 3421 | 100 | 40 | 0.223 | 0.022 | 10.041 | 26.39 | < 0.001 | [0.177, 0.268] | 73.1 |
| 3422 | 100 | 40 | 0.222 | 0.022 | 9.993 | 26.42 | < 0.001 | [0.177, 0.268] | 73.2 |
| 3431 | 100 | 40 | 0.223 | 0.022 | 10.082 | 26.33 | < 0.001 | [0.178, 0.269] | 73.0 |

Table S6. Leave-one-out results: Effect size (*continued*)
| Left-out<br>ES ID | $k$ | Clusters | Hedges' $g$ | $SE$ | $t$ | $df$ | $p$ | 95% CI | $I^2$ (%) |
| --- | --- | --- | --- | --- | --- | --- | --- | --- | --- |
| 3432 | 100 | 40 | 0.223 | 0.022 | 9.990 | 26.55 | < 0.001 | [0.177, 0.268] | 73.2 |
| 3441 | 100 | 40 | 0.221 | 0.022 | 9.890 | 26.36 | < 0.001 | [0.175, 0.267] | 73.4 |
| 3442 | 100 | 40 | 0.223 | 0.022 | 10.078 | 26.33 | < 0.001 | [0.178, 0.269] | 73.3 |
| 3443 | 100 | 40 | 0.223 | 0.022 | 10.041 | 26.34 | < 0.001 | [0.177, 0.268] | 73.3 |
| 3451 | 100 | 40 | 0.223 | 0.022 | 10.085 | 26.33 | < 0.001 | [0.178, 0.269] | 73.2 |
| 3452 | 100 | 40 | 0.222 | 0.022 | 9.959 | 26.37 | < 0.001 | [0.176, 0.268] | 73.4 |
| 3453 | 100 | 40 | 0.223 | 0.022 | 10.020 | 26.36 | < 0.001 | [0.177, 0.268] | 73.3 |
| 3461 | 100 | 40 | 0.222 | 0.022 | 9.981 | 26.37 | < 0.001 | [0.176, 0.268] | 73.3 |
| 3462 | 100 | 40 | 0.222 | 0.022 | 10.003 | 26.36 | < 0.001 | [0.177, 0.268] | 73.3 |
| 3463 | 100 | 40 | 0.222 | 0.022 | 9.994 | 26.37 | < 0.001 | [0.177, 0.268] | 73.3 |
| 3511 | 100 | 39 | 0.220 | 0.022 | 9.938 | 25.88 | < 0.001 | [0.174, 0.265] | 72.9 |
| 3711 | 100 | 40 | 0.222 | 0.023 | 9.828 | 25.97 | < 0.001 | [0.176, 0.269] | 73.5 |
| 3722 | 100 | 40 | 0.223 | 0.023 | 9.865 | 25.96 | < 0.001 | [0.177, 0.270] | 73.5 |
| 3811 | 100 | 40 | 0.221 | 0.022 | 9.869 | 25.91 | < 0.001 | [0.175, 0.267] | 73.4 |
| 3821 | 100 | 40 | 0.222 | 0.022 | 9.872 | 25.91 | < 0.001 | [0.175, 0.268] | 73.5 |
| 3931 | 100 | 40 | 0.225 | 0.023 | 9.910 | 26.37 | < 0.001 | [0.178, 0.271] | 72.9 |
| 3941 | 100 | 40 | 0.219 | 0.023 | 9.542 | 26.33 | < 0.001 | [0.172, 0.266] | 72.4 |
| 4011 | 100 | 40 | 0.226 | 0.021 | 10.613 | 25.96 | < 0.001 | [0.183, 0.270] | 72.0 |
| 4021 | 100 | 40 | 0.227 | 0.021 | 10.639 | 25.92 | < 0.001 | [0.183, 0.270] | 71.3 |
| 4111 | 100 | 40 | 0.222 | 0.023 | 9.818 | 26.02 | < 0.001 | [0.176, 0.269] | 73.0 |
| 4121 | 100 | 40 | 0.223 | 0.023 | 9.828 | 26.02 | < 0.001 | [0.176, 0.269] | 73.1 |
| 4212 | 100 | 39 | 0.223 | 0.023 | 9.884 | 25.53 | < 0.001 | [0.177, 0.270] | 73.5 |

**Table S7.** Leave-one-out results: Experiment.

| Left-out<br>Exp. ID | $k$ | Clusters | Hedges' $g$ | $SE$ | $t$ | $df$ | $p$ | 95% CI | $I^2$ (%) |
| --- | --- | --- | --- | --- | --- | --- | --- | --- | --- |
| 11 | 98 | 39 | 0.220 | 0.022 | 9.833 | 25.49 | < 0.001 | [0.174, 0.266] | 73.3 |
| 21 | 99 | 39 | 0.219 | 0.022 | 9.878 | 25.61 | < 0.001 | [0.174, 0.265] | 73.1 |
| 31 | 100 | 40 | 0.222 | 0.023 | 9.863 | 25.92 | < 0.001 | [0.176, 0.269] | 73.6 |
| 32 | 100 | 40 | 0.221 | 0.022 | 9.850 | 25.92 | < 0.001 | [0.175, 0.267] | 73.4 |
| 41 | 100 | 39 | 0.223 | 0.022 | 9.982 | 25.84 | < 0.001 | [0.177, 0.269] | 73.5 |
| 51 | 100 | 39 | 0.216 | 0.022 | 9.930 | 25.34 | < 0.001 | [0.172, 0.261] | 71.1 |
| 61 | 100 | 39 | 0.223 | 0.022 | 9.933 | 25.72 | < 0.001 | [0.177, 0.269] | 73.6 |
| 71 | 100 | 39 | 0.222 | 0.022 | 9.957 | 25.91 | < 0.001 | [0.177, 0.268] | 73.5 |
| 81 | 99 | 40 | 0.224 | 0.022 | 10.008 | 26.00 | < 0.001 | [0.178, 0.271] | 73.2 |
| 82 | 99 | 40 | 0.225 | 0.022 | 10.017 | 26.00 | < 0.001 | [0.178, 0.271] | 73.2 |
| 91 | 99 | 40 | 0.223 | 0.023 | 9.823 | 26.00 | < 0.001 | [0.177, 0.270] | 73.5 |
| 94 | 99 | 40 | 0.220 | 0.022 | 9.780 | 25.96 | < 0.001 | [0.174, 0.266] | 72.9 |
| 101 | 100 | 40 | 0.223 | 0.023 | 9.874 | 25.91 | < 0.001 | [0.176, 0.269] | 73.6 |
| 104 | 100 | 40 | 0.225 | 0.022 | 10.027 | 25.88 | < 0.001 | [0.179, 0.271] | 73.2 |
| 111 | 99 | 40 | 0.220 | 0.022 | 9.885 | 26.52 | < 0.001 | [0.174, 0.265] | 73.2 |
| 121 | 99 | 40 | 0.222 | 0.022 | 9.906 | 26.57 | < 0.001 | [0.176, 0.268] | 73.7 |
| 131 | 100 | 39 | 0.225 | 0.023 | 9.989 | 25.43 | < 0.001 | [0.179, 0.272] | 72.4 |
| 141 | 100 | 39 | 0.223 | 0.023 | 9.895 | 25.58 | < 0.001 | [0.177, 0.270] | 73.6 |

Table S7. Leave-one-out results: Experiment (*continued*)
| Left-out<br>Exp. ID | $k$ | Clusters | Hedges' $g$ | $SE$ | $t$ | $df$ | $p$ | 95% CI | $I^2$ (%) |
| --- | --- | --- | --- | --- | --- | --- | --- | --- | --- |
| 151 | 100 | 40 | 0.223 | 0.023 | 9.907 | 25.96 | < 0.001 | [0.177, 0.270] | 73.5 |
| 152 | 100 | 40 | 0.224 | 0.023 | 9.966 | 25.95 | < 0.001 | [0.178, 0.271] | 73.3 |
| 161 | 99 | 40 | 0.218 | 0.022 | 10.035 | 25.85 | < 0.001 | [0.173, 0.262] | 72.3 |
| 162 | 99 | 40 | 0.221 | 0.022 | 9.961 | 25.92 | < 0.001 | [0.175, 0.267] | 73.6 |
| 171 | 99 | 40 | 0.225 | 0.022 | 10.118 | 25.83 | < 0.001 | [0.179, 0.271] | 73.3 |
| 172 | 99 | 40 | 0.224 | 0.022 | 10.052 | 25.94 | < 0.001 | [0.178, 0.270] | 73.7 |
| 181 | 99 | 39 | 0.227 | 0.022 | 10.201 | 25.42 | < 0.001 | [0.181, 0.272] | 72.2 |
| 191 | 100 | 39 | 0.224 | 0.022 | 9.955 | 25.70 | < 0.001 | [0.177, 0.270] | 73.5 |
| 201 | 100 | 39 | 0.217 | 0.022 | 10.007 | 25.70 | < 0.001 | [0.172, 0.261] | 71.6 |
| 211 | 99 | 40 | 0.221 | 0.022 | 9.843 | 26.66 | < 0.001 | [0.175, 0.267] | 73.3 |
| 213 | 99 | 40 | 0.225 | 0.023 | 9.740 | 26.64 | < 0.001 | [0.178, 0.272] | 73.3 |
| 221 | 99 | 40 | 0.226 | 0.022 | 10.456 | 25.90 | < 0.001 | [0.182, 0.270] | 72.7 |
| 224 | 99 | 40 | 0.226 | 0.022 | 10.462 | 25.90 | < 0.001 | [0.182, 0.270] | 72.7 |
| 231 | 99 | 39 | 0.225 | 0.023 | 10.003 | 25.44 | < 0.001 | [0.179, 0.272] | 72.8 |
| 241 | 99 | 40 | 0.224 | 0.023 | 9.876 | 25.96 | < 0.001 | [0.177, 0.270] | 73.7 |
| 242 | 99 | 40 | 0.221 | 0.023 | 9.787 | 25.90 | < 0.001 | [0.175, 0.267] | 73.5 |
| 251 | 100 | 40 | 0.223 | 0.023 | 9.854 | 25.91 | < 0.001 | [0.177, 0.270] | 73.5 |
| 252 | 100 | 40 | 0.221 | 0.022 | 9.821 | 25.94 | < 0.001 | [0.175, 0.267] | 73.3 |
| 261 | 100 | 40 | 0.223 | 0.022 | 10.004 | 26.06 | < 0.001 | [0.177, 0.269] | 73.4 |
| 262 | 100 | 40 | 0.223 | 0.022 | 10.011 | 26.04 | < 0.001 | [0.177, 0.269] | 73.4 |
| 263 | 100 | 40 | 0.222 | 0.022 | 9.975 | 26.06 | < 0.001 | [0.176, 0.268] | 73.4 |
| 271 | 100 | 39 | 0.225 | 0.022 | 10.069 | 25.66 | < 0.001 | [0.179, 0.271] | 73.1 |
| 281 | 100 | 40 | 0.219 | 0.022 | 9.999 | 25.85 | < 0.001 | [0.174, 0.264] | 72.8 |
| 282 | 100 | 40 | 0.221 | 0.022 | 9.978 | 25.87 | < 0.001 | [0.175, 0.266] | 73.3 |
| 291 | 100 | 39 | 0.216 | 0.021 | 10.056 | 25.64 | < 0.001 | [0.172, 0.260] | 70.9 |
| 301 | 100 | 39 | 0.218 | 0.022 | 9.889 | 25.51 | < 0.001 | [0.173, 0.263] | 72.3 |
| 311 | 100 | 40 | 0.223 | 0.023 | 9.906 | 25.98 | < 0.001 | [0.177, 0.270] | 73.5 |
| 318 | 100 | 40 | 0.224 | 0.023 | 9.930 | 25.93 | < 0.001 | [0.178, 0.270] | 73.3 |
| 321 | 100 | 40 | 0.223 | 0.022 | 10.016 | 26.16 | < 0.001 | [0.177, 0.268] | 73.3 |
| 322 | 100 | 40 | 0.222 | 0.022 | 10.014 | 26.16 | < 0.001 | [0.177, 0.268] | 73.3 |
| 323 | 100 | 40 | 0.222 | 0.022 | 10.012 | 26.16 | < 0.001 | [0.177, 0.268] | 73.3 |
| 331 | 100 | 39 | 0.223 | 0.022 | 10.001 | 26.07 | < 0.001 | [0.177, 0.268] | 73.4 |
| 341 | 99 | 40 | 0.219 | 0.023 | 9.474 | 27.13 | < 0.001 | [0.172, 0.266] | 72.9 |
| 342 | 99 | 40 | 0.224 | 0.022 | 9.984 | 27.55 | < 0.001 | [0.178, 0.270] | 73.5 |
| 343 | 99 | 40 | 0.225 | 0.022 | 10.087 | 27.46 | < 0.001 | [0.180, 0.271] | 73.1 |
| 344 | 98 | 40 | 0.223 | 0.023 | 9.910 | 27.47 | < 0.001 | [0.177, 0.269] | 73.9 |
| 345 | 98 | 40 | 0.224 | 0.022 | 10.029 | 27.38 | < 0.001 | [0.178, 0.270] | 73.8 |
| 346 | 98 | 40 | 0.222 | 0.023 | 9.830 | 27.32 | < 0.001 | [0.176, 0.268] | 74.0 |
| 351 | 100 | 39 | 0.220 | 0.022 | 9.938 | 25.88 | < 0.001 | [0.174, 0.265] | 72.9 |
| 361 | 100 | 40 | 0.219 | 0.022 | 10.145 | 25.90 | < 0.001 | [0.175, 0.263] | 72.8 |
| 363 | 100 | 40 | 0.219 | 0.022 | 10.128 | 25.77 | < 0.001 | [0.174, 0.263] | 72.5 |
| 371 | 100 | 40 | 0.222 | 0.023 | 9.828 | 25.97 | < 0.001 | [0.176, 0.269] | 73.5 |
| 372 | 100 | 40 | 0.223 | 0.023 | 9.865 | 25.96 | < 0.001 | [0.177, 0.270] | 73.5 |
| 381 | 100 | 40 | 0.221 | 0.022 | 9.869 | 25.91 | < 0.001 | [0.175, 0.267] | 73.4 |
| 382 | 100 | 40 | 0.222 | 0.022 | 9.872 | 25.91 | < 0.001 | [0.175, 0.268] | 73.5 |
| 391 | 100 | 40 | 0.225 | 0.023 | 9.968 | 26.37 | < 0.001 | [0.178, 0.271] | 73.3 |

Table S7. Leave-one-out results: Experiment (*continued*)
| Left-out<br>Exp. ID | $k$ | Clusters | Hedges' $g$ | $SE$ | $t$ | $df$ | $p$ | 95% CI | $I^2$ (%) |
| --- | --- | --- | --- | --- | --- | --- | --- | --- | --- |
| 393 | 100 | 40 | 0.225 | 0.023 | 9.910 | 26.37 | < 0.001 | [0.178, 0.271] | 72.9 |
| 394 | 100 | 40 | 0.219 | 0.023 | 9.542 | 26.33 | < 0.001 | [0.172, 0.266] | 72.4 |
| 401 | 100 | 40 | 0.226 | 0.021 | 10.613 | 25.96 | < 0.001 | [0.183, 0.270] | 72.0 |
| 402 | 100 | 40 | 0.227 | 0.021 | 10.639 | 25.92 | < 0.001 | [0.183, 0.270] | 71.3 |
| 411 | 100 | 40 | 0.222 | 0.023 | 9.818 | 26.02 | < 0.001 | [0.176, 0.269] | 73.0 |
| 412 | 100 | 40 | 0.223 | 0.023 | 9.828 | 26.02 | < 0.001 | [0.176, 0.269] | 73.1 |
| 421 | 100 | 39 | 0.223 | 0.023 | 9.884 | 25.53 | < 0.001 | [0.177, 0.270] | 73.5 |

**Table S8.**
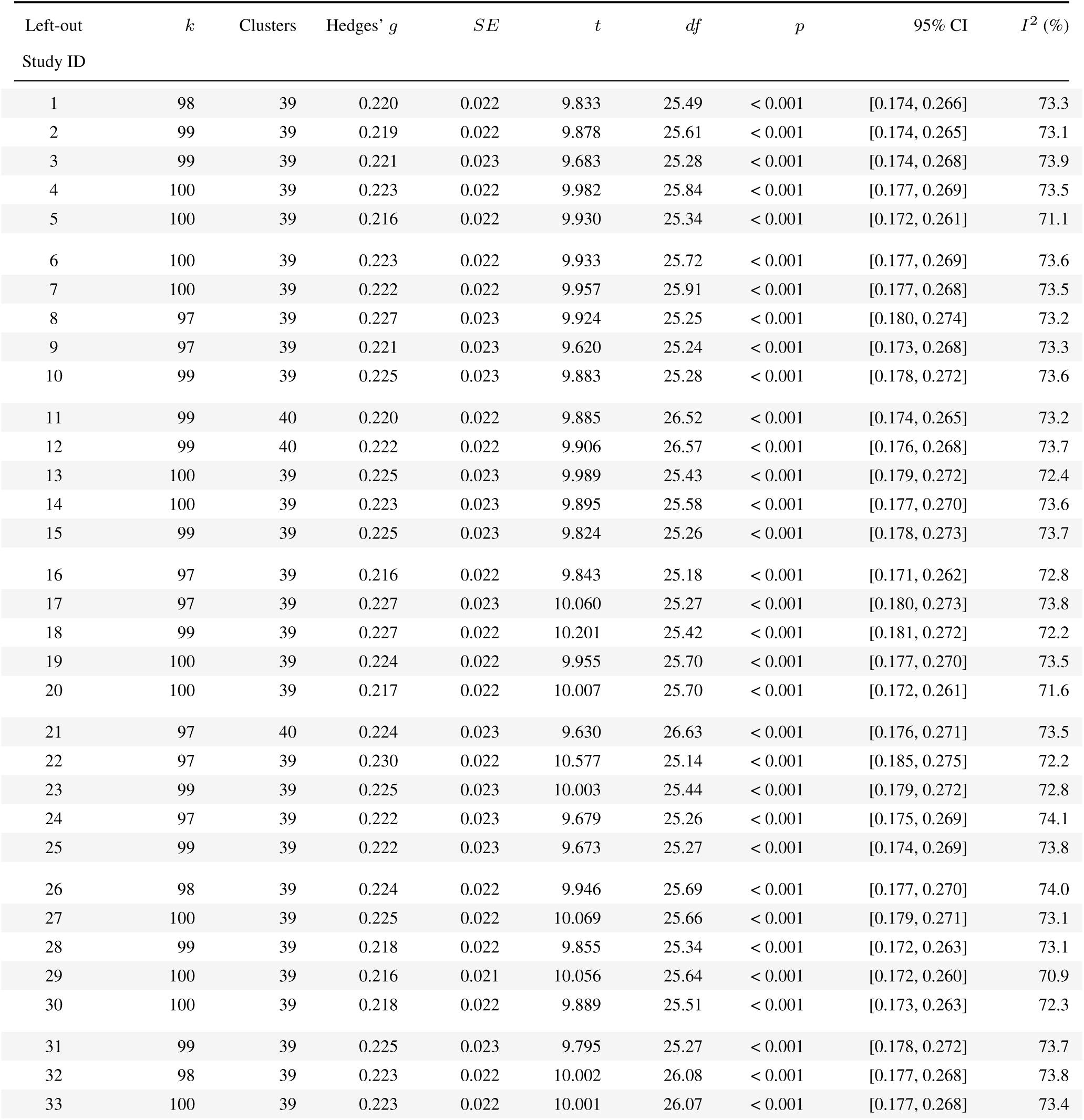
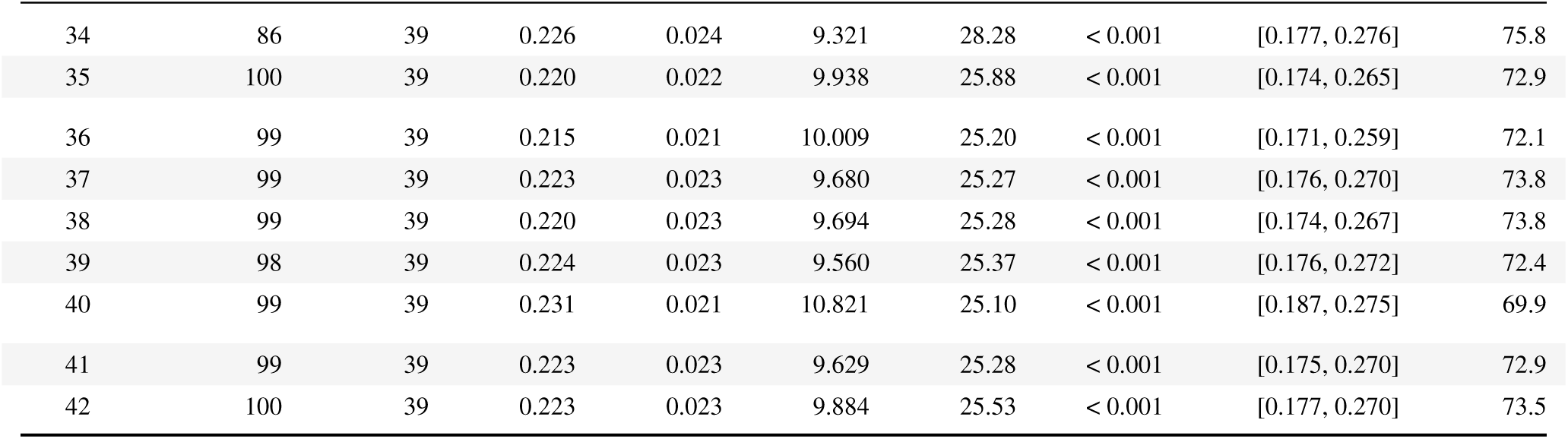
Leave-one-out results: Study.

## References

[1] Thammasitboon, S. & Cutrer, W. B. Diagnostic decision-making and strategies to improve diagnosis. Curr Probl Pediatr Adolesc Health Care 43, 232–241 (2013).

[2] Newman-Toker, D. E. et al. Burden of serious harms from diagnostic error in the USA. BMJ Qual Saf 33, 109–120 (2024).

[3] Feuerriegel, S., Hartmann, J., Janiesch, C. & Zschech, P. Generative AI. Bus Inf Syst Eng 66, 111–126 (2024).

[4] Topol, E. J. Toward the eradication of medical diagnostic errors. Science 383, eadn9602 (2024).

[5] Kanjee, Z., Crowe, B. & Rodman, A. Accuracy of a generative artificial intelligence model in a complex diagnostic challenge. JAMA 330, 78–80 (2023).

[6] Williams, C. Y. K., Miao, B. Y., Kornblith, A. E. & Butte, A. J. Evaluating the use of large language models to provide clinical recommendations in the emergency department. Nat Commun 15, 8236 (2024).

[7] Goh, E. et al. GPT-4 assistance for improvement of physician performance on patient care tasks: a randomized controlled trial. Nat Med 31, 1233–1238 (2025).

[8] Takita, H. et al. A systematic review and meta-analysis of diagnostic performance comparison between generative AI and physicians. npj Digit Med 8, 175 (2025).

[9] Liu, F. et al. MetaGP: a generative foundation model integrating electronic health records and multimodal imaging for addressing unmet clinical needs. Cell Rep Med 6, 102056 (2025).

[10] Schmidgall, S. et al. Evaluation and mitigation of cognitive biases in medical language models. npj Digit Med 7, 295 (2024).

[11] Ke, Y. et al. Mitigating cognitive biases in clinical decision-making through multi-agent conversations using large language models: simulation study. J Med Internet Res 26, e59439 (2024).

[12] Zöller, N. et al. Human–AI collectives most accurately diagnose clinical vignettes. Proc Natl Acad Sci USA 122, e2426153122 (2025).

[13] Zhou, S. et al. Large language models for disease diagnosis: a scoping review. npj Artif Intell 1, 9 (2025).

[14] Eichenberger, A., Thielke, S. & Van Buskirk, A. A case of bromism influenced by use of artificial intelligence. Ann Intern Med Clin Cases 4, e241260 (2025).

[15] Kim, S. H. et al. Human-AI collaboration in large language model-assisted brain MRI differential diagnosis: a usability study. Eur Radiol 35, 5252–5263 (2025).

[16] Wang, G. et al. Human–large language model collaboration in clinical medicine: a systematic review and meta-analysis. npj Digit Med 9, 195 (2026).

[17] Chen, M. et al. Independent and collaborative performance of large language models and health-care professionals in diagnosis and triage. npj Digit Med 9, 222 (2026).

[18] Spitzer, P. et al. The effect of medical explanations from large language models on diagnostic accuracy in radiology. npj Digit Med **9**, 333 (2026).

[19] Atakır, K., Isın, K., Tas, A. & Önder, H. Diagnostic accuracy and consistency of ChatGPT-4o in radiology: influence of image, clinical data, and answer options on performance. Diagn Interv Radiol (2025). Advance online publication.

[20] Çamur, E., Cesur, T., Günes, Y. C., Gökhan, M. B. & Ökten, R. S. Prospective quantitative analysis of hyperparameter and input optimization in GPT-5: comparative contribution to radiologist performance in abdominal radiology. Diagn Interv Radiol (2026). Advance online publication.

[21] Cesur, T., Günes, Y. C., Çamur, E. & Daglı, M. Empowering radiologists with ChatGPT-4o: Comparative evaluation of large language models and radiologists in cardiac cases. J Thorac Imaging 40, e0846 (2025).

[22] Chen, X. et al. Grounding large language models in clinical diagnostics. Nat Commun 17, 4401 (2026).

[23] Dezoteux, F. et al. AI-assisted dermatologic diagnosis using a large language model. J Am Acad Dermatol 94, 976–978 (2026).

[24] Goh, E. et al. Large language model influence on diagnostic reasoning: a randomized clinical trial. JAMA Netw Open 7, e2440969 (2024).

[25] Healy, J., Kossoff, J., Lee, M. & Hasford, C. Human–AI collaboration in clinical reasoning: a UK replication and interaction analysis. Diagnosis (2026). Advance online publication.

[26] Horiuchi, D. et al. ChatGPT’s diagnostic performance based on textual vs. visual information compared to radiologists’ diagnostic performance in musculoskeletal radiology. Eur Radiol 35, 506–516 (2025).

[27] Ji, X. et al. Early diagnosis of axial spondyloarthritis in primary care using multi-agent systems. npj Digit Med 9, 185 (2026).

[28] Jin, D.-D. et al. ChatGPT-4V prompt: A tool to enhance junior radiologists’ diagnostic capabilities in cystic renal masses to senior-level accuracy. Comput Struct Biotechnol J 32, 34–47 (2026).

[29] Kim, S. H. et al. Boosting LLM-assisted diagnosis: 10-minute LLM tutorial elevates radiology residents’ performance in brain MRI interpretation. Neuroradiology 67, 2069–2081 (2025).

30. [30] Kowshik, S. S. et al. Domain-adapted language model using reinforcement learning for various dementias. Preprint at medRxiv 10.64898/2026.03.17.26348154 (2026).

[31] Le Guellec, B. et al. Comparison between multimodal foundation models and radiologists for the diagnosis of challenging neuroradiology cases with text and images. Diagn Interv Imaging 106, 345–352 (2025).

[32] Liu, X. et al. A generalist medical language model for disease diagnosis assistance. Nat Med 31, 932–942 (2025).

[33] Long, H. et al. ChatGPT-4o assists emergency physicians in enhancing diagnostic accuracy for fever of unknown origin: retrospective analysis. BMC Emerg Med 26, 45 (2026).

[34] McDuff, D. et al. Towards accurate differential diagnosis with large language models. Nature 642, 451–457 (2025).

[35] Mukherjee, P. et al. Evaluation of GPT large language model performance on RSNA 2023 case of the day questions. Radiology 313, e240609 (2024).

[36] Oon, M. L., Syn, N. L., Tan, C. L., Tan, K.-B. & Ng, S.-B. Bridging bytes and biopsies: A comparative analysis of ChatGPT and histopathologists in pathology diagnosis and collaborative potential. Histopathology 84, 601–613 (2024).

[37] Qazi, I. A. et al. Large language model diagnostic assistance for physicians in a lower-middle-income country: a randomized controlled trial. Nat Health 1, 198–205 (2026).

[38] Schramm, S. et al. Performing best when needed least: Reader experience shapes accuracy gains in large language model–assisted brain MRI differential diagnosis. Radiology 319, e253477 (2026).

[39] Sheng, L. et al. Large language models for diagnosing focal liver lesions from CT/MRI reports: a comparative study with radiologists. Liver Int 45, e70115 (2025).

[40] Siepmann, R. et al. The virtual reference radiologist: comprehensive AI assistance for clinical image reading and interpretation. Eur Radiol 34, 6652–6666 (2024).

[41] Song, J. et al. Human-in-the-loop large language model–augmented diagnostic reasoning in thoracic imaging: impact of radiologic expertise. AJR Am J Roentgenol (2026). Advance online publication.

[42] Song, C. W. et al. Impact of large language model assistance on radiologists’ diagnostic performance for brain tumors by experience level. J Clin Med 15, 1673 (2026).

[43] Ver Berne, J., Baseri Saadi, S., Silva da Andrade-Bortoletto, M. F., Fontenele, R. & Jacobs, R. Impact of AI-assisted decision support on radiological diagnosis of jawbone lesions. Clin Oral Investig 30, 61 (2026).

[44] Wang, Y. et al. Ultrasound-based deep learning model as an assistant improves the diagnosis of ovarian tumors: a multicenter study. Insights Imaging 16, 221 (2025).

[45] Wang, R. et al. A domain-adapted large language model to support clinicians in psychiatric clinical practice. Nat Mach Intell 8, 690–707 (2026).

[46] Wang, M.-L. et al. Evaluation of large language models for diagnostic impression generation from brain MRI report findings: a multicenter benchmark and reader study. npj Digit Med 9, 187 (2026).

[47] Wang, M. et al. Enhancing diagnostic accuracy in rare and common fundus diseases with a knowledge-rich vision-language model. Nat Commun 16, 5528 (2025).

[48] Wang, Z., Guo, R., Sun, P., Qian, L. & Hu, X. Enhancing diagnostic accuracy and efficiency with GPT-4-generated structured reports: a comprehensive study. J Med Biol Eng 44, 144–153 (2024).

[49] Weuthen, F. A., Otte, N., Krabbe, H., Kraus, T. & Krabbe, J. Comparison of ChatGPT and internet research for clinical research and decision-making in occupational medicine: randomized controlled trial. JMIR Form Res 9, e63857 (2025).

[50] Wu, X., Huang, Y. & He, Q. A large language model improves clinicians’ diagnostic performance in complex critical illness cases. Crit Care 29, 230 (2025).

[51] Wu, Y. et al. An eyecare foundation model for clinical assistance: a randomized controlled trial. Nat Med 31, 3404–3413 (2025).

[52] Xu, N. et al. Predicting invasiveness of lung adenocarcinoma from chest CT with few-shot vision-language ternary classification model. npj Digit Med 9, 60 (2026).

[53] Yan, S. et al. A multimodal vision foundation model for clinical dermatology. Nat Med 31, 2691–2702 (2025).

[54] Yao, J. et al. Multimodal GPT model for assisting thyroid nodule diagnosis and management. npj Digit Med **8**, 245 (2025).

[55] Zhang, S. et al. A novel approach to ovarian cancer diagnosis via CT imaging: GPT-4o-driven automated feature recognition and validation in clinical settings. Ann Surg Oncol 33, 6618–6628 (2026).

[56] Zhou, S. et al. Uncertainty-aware large language models for explainable disease diagnosis. npj Digit Med **8**, 690 (2025).

57. Zhou, S., et al. HeartAgent: An autonomous agent system for explainable differential diagnosis in cardiology. Preprint at arXiv 10.48550/arXiv.2603.10764 (2026).

58. Zhou, S., et al. EpiScreen: Early epilepsy detection from electronic health records with large language models. Preprint at arXiv 10.48550/arXiv.2603.28698 (2026).

[59] Egger, M., Davey Smith, G., Schneider, M. & Minder, C. Bias in meta-analysis detected by a simple, graphical test. BMJ 315, 629–634 (1997).

60. Gliadkovskaya, A. Some doctors are using ChatGPT to assist with clinical decisions. Is it safe? (2024). URL https://www.fiercehealthcare.com/special-reports/some-doctors-are-using-public-generative-ai-tools-chatgpt-clinical-decisions-it. Accessed on August 26, 2026.

[61] Meyer, A., Riese, J. & Streichert, T. Comparison of the performance of GPT-3.5 and GPT-4 with that of medical students on the written German medical licensing examination: observational study. JMIR Med Educ 10, e50965 (2024).

[62] Masanneck, L. et al. Triage performance across large language models, ChatGPT, and untrained doctors in emergency medicine: comparative study. J Med Internet Res 26, e53297 (2024).

[63] Sousa, D., Barbosa, G., Rocha, C. & Oliveira, D. Performance of large language models in supporting medical diagnosis and treatment. Preprint at arXiv 10.48550/arXiv.2504.10405 (2025).

[64] Hirosawa, T. et al. ChatGPT-generated differential diagnosis lists for complex case–derived clinical vignettes: diagnostic accuracy evaluation. JMIR Med Inform 11, e48808 (2023).

[65] Vered, M., Livni, T., Howe, P. D. L., Miller, T. & Sonenberg, L. The effects of explanations on automation bias. Artif Intell 322, 103952 (2023).

[66] Everett, S. S. et al. From tool to teammate in a randomized controlled trial of clinician-AI collaborative workflows for diagnosis. npj Digit Med **9**, 409 (2026).

[67] Fogliato, R. et al. Who goes first? Influences of human-AI workflow on decision making in clinical imaging. FAccT ’22: Proceedings of the 2022 ACM Conference on Fairness, Accountability, and Transparency, 1362–1374 (Association for Computing Machinery, New York, NY, USA, 2022). URL 10.1145/3531146.3533193.

[68] Prucker, P. et al. Performance of open-source and proprietary large language models in generating patient-friendly radiology chest CT reports. Clin Imaging 125, 110557 (2025).

[69] Xie, Q. et al. Medical foundation large language models for comprehensive text analysis and beyond. npj Digit Med 8, 141 (2025).

70. Bandyopadhyay, D., Bhattacharjee, S. & Ekbal, A. Thinking machines: A survey of LLM based reasoning strategies. Preprint at arXiv 10.48550/arXiv.2503.10814 (2025).

[71] Yoon, J. H., Pinsky, M. R. & Clermont, G. Artificial intelligence in critical care medicine. Crit Care 26, 75 (2022).

[72] Bedi, S. et al. Testing and evaluation of health care applications of large language models: a systematic review. JAMA 333, 319–328 (2025).

73. Kühl, N., et al. A human-centered approach to designing large language models in medicine. Preprint at SSRN 10.2139/ssrn.5569098 (2025).

[74] Omar, M. et al. Evaluating and addressing demographic disparities in medical large language models: a systematic review. Int J Equity Health 24, 57 (2025).

[75] Omiye, J. A., Lester, J. C., Spichak, S., Rotemberg, V. & Daneshjou, R. Large language models propagate race-based medicine. npj Digit Med **6**, 195 (2023).

[76] Zack, T. et al. Assessing the potential of GPT-4 to perpetuate racial and gender biases in health care: a model evaluation study. Lancet Digit Health 6, e12–e22 (2024).

[77] Page, M. J. et al. The PRISMA 2020 statement: an updated guideline for reporting systematic reviews. BMJ 372, n71 (2021).

78. Tretow, I., Feuerriegel, S., Schwebel, M., Treffers, T. & Welpe, I. M. The effect of LLM assistance on diagnostic accuracy: a meta-analysis. PROSPERO 2025 CRD420251067623. https://www.crd.york.ac.uk/PROSPERO/view/CRD420251067623 (2025). Accessed on August 11, 2025.

[79] Brown, T. et al. Language models are few-shot learners. Advances in Neural Information Processing Systems, Vol. 33, 1877–1901 (Curran Associates, Inc., 2020). URL https://proceedings.neurips.cc/paper_files/paper/2020/file/1457c0d6bfcb4967418bfb8ac142f64a-Paper.pdf.

80. Higgins, J. P. et al. Cochrane Handbook for Systematic Reviews of Interventions version 6.5 (updated August 2024) (Cochrane, 2024). URL https://www.cochrane.org/handbook. Accessed on August 25, 2026.

[81] Vaccaro, M., Almaatouq, A. & Malone, T. When combinations of humans and AI are useful: A systematic review and meta-analysis. Nat Hum Behav 8, 2293–2303 (2024).

[82] Ouzzani, M., Hammady, H., Fedorowicz, Z. & Elmagarmid, A. Rayyan—a web and mobile app for systematic reviews. Syst Rev 5, 210 (2016).

[83] Wolff, R. F. et al. PROBAST: a tool to assess the risk of bias and applicability of prediction model studies. Ann Intern Med 170, 51–58 (2019).

84. [84] Hedges, L. V. & Olkin, I. Statistical Methods for Meta-Analysis (Academic Press, Orlando, FL, 1985). Ch. 5, pp. 75–106.

[85] Veroniki, A. A. et al. Methods to estimate the between-study variance and its uncertainty in meta-analysis. Res Synth Methods 7, 55–79 (2016).

[86] Van den Noortgate, W., López-López, J. A., Marín-Martínez, F. & Sánchez-Meca, J. Meta-analysis of multiple outcomes: a multilevel approach. Behav Res Methods 47, 1274–1294 (2015).

[87] Tipton, E. & Pustejovsky, J. E. Small-sample adjustments for tests of moderators and model fit using robust variance estimation in meta-regression. J Educ Behav Stat 40, 604–634 (2015).

[88] Tipton, E. Small sample adjustments for robust variance estimation with meta-regression. Psychol Methods 20, 375–393 (2015).

[89] Higgins, J. P. T. & Thompson, S. G. Quantifying heterogeneity in a meta-analysis. Stat Med 21, 1539–1558 (2002). URL https://onlinelibrary.wiley.com/doi/abs/10.1002/sim.1186.

90. R Core Team. *R: A language and environment for statistical computing*. R Foundation for Statistical Computing, Vienna, Austria (2025). URL https://www.R-project.org. Accessed on August 25, 2026.

[91] Viechtbauer, W. Conducting meta-analyses in R with the metafor package. J Stat Softw 36, 1–48 (2010).

[92] Pustejovsky, J. E. clubSandwich: Cluster-Robust (Sandwich) Variance Estimators with Small-Sample Corrections (2026). URL https://CRAN.R-project.org/package=clubSandwich. R pack-age version 0.7.0. Accessed on August 25, 2026.

## References (Appendix)

[1] Sutton, R. T. et al. An overview of clinical decision support systems: benefits, risks, and strategies for success. npj Digit Med **3**, 17 (2020).

[2] Thirunavukarasu, A. J. et al. Large language models in medicine. Nat Med 29, 1930–1940 (2023).

[3] Singhal, K. et al. Large language models encode clinical knowledge. Nature 620, 172–180 (2023).

[4] Takita, H. et al. A systematic review and meta-analysis of diagnostic performance comparison between generative AI and physicians. npj Digit Med **8**, 175 (2025).

[5] Siepmann, R. et al. The virtual reference radiologist: comprehensive AI assistance for clinical image reading and interpretation. Eur Radiol 34, 6652–6666 (2024).

[6] McDuff, D. et al. Towards accurate differential diagnosis with large language models. Nature 642, 451–457 (2025).

[7] Spitzer, P. et al. The effect of medical explanations from large language models on diagnostic accuracy in radiology. npj Digit Med **9**, 333 (2026).

[8] Cesur, T., Günes, Y. C., Çamur, E. & Daglı, M. Empowering radiologists with ChatGPT-4o: Comparative evaluation of large language models and radiologists in cardiac cases. J Thorac Imaging 40, e0846 (2025).

[9] Mukherjee, P. et al. Evaluation of GPT large language model performance on RSNA 2023 case of the day questions. Radiology 313, e240609 (2024).

[10] Wang, Y. et al. Ultrasound-based deep learning model as an assistant improves the diagnosis of ovarian tumors: a multicenter study. Insights Imaging 16, 221 (2025).

[11] Sheng, L. et al. Large language models for diagnosing focal liver lesions from CT/MRI reports: a comparative study with radiologists. Liver Int 45, e70115 (2025).

[12] Viechtbauer, W. Conducting meta-analyses in R with the metafor package. J Stat Softw 36, 1–48 (2010).

[13] Hedges, L. V. & Olkin, I. Statistical Methods for Meta-Analysis (Academic Press, Orlando, FL, 1985). Ch. 5, pp. 75–106.

[14] Wang, R. et al. A domain-adapted large language model to support clinicians in psychiatric clinical practice. Nat Mach Intell 8, 690–707 (2026).

[15] Tipton, E. & Pustejovsky, J. E. Small-sample adjustments for tests of moderators and model fit using robust variance estimation in meta-regression. J Educ Behav Stat 40, 604–634 (2015).

[16] Pustejovsky, J. E. & Rodgers, M. A. Testing for funnel plot asymmetry of standardized mean differences. Res Synth Methods 10, 57–71 (2019).

[17] Kim, S. H. et al. Boosting LLM-assisted diagnosis: 10-minute LLM tutorial elevates radiology residents’ performance in brain MRI interpretation. Neuroradiology 67, 2069–2081 (2025).

[18] Kim, S. H. et al. Human-AI collaboration in large language model-assisted brain MRI differential diagnosis: a usability study. Eur Radiol 35, 5252–5263 (2025).

